# Development and validation of polygenic scores for within-family prediction of disease risks

**DOI:** 10.1101/2025.08.06.25333145

**Authors:** Spencer Moore, Ivan Davidson, Jonathan Anomaly, Jeremiah H. Li, Mohammad Ahangari, Lauren Moissiy, Michael Christensen, Alexander Strudwick Young, David Stern, Tobias Wolfram

**Affiliations:** Herasight Research, USA; Human Genetics Department, UCLA David Geffen School of Medicine, Los Angeles, CA, US

## Abstract

The clinical implementation of polygenic scores (PGSs) for disease risk prediction, particularly in reproductive health applications, requires rigorous validation. Here, we develop seventeen disease PGSs by conducting large-scale GWAS meta-analyses, and we validate our scores in out-of-sample prediction analyses. We achieve state-of-the-art predictive performance, consistently matching or outperforming academic and commercial benchmarks, with liability *R*^2^ reaching up to 0.21 (type 2 diabetes). The performance of a PGS for embryo screening depends on its predictive ability within-family, which can be lower than its prediction ability among unrelated individuals. However, very few disease PGSs have been tested within-family. We perform systematic within-family validation of our disease PGSs, finding no decrease in predictive performance within-family for 16 of 17 scores. PGS performance typically declines with genetic distance from training data, an effect that needs to be accounted for to give properly calibrated predictions across ancestries. We perform extensive calibration of our scores’ performance across different ancestries, finding improved cross-ancestry performance compared to previous approaches, especially in African and East Asian populations. This is likely due to the fact our scores are constructed using a method that incorporates functional genomic annotations on more than 7 million variants, enabling a degree of fine-mapping of causal variants shared across ancestries. We illustrate clinical utility through examining the risk reduction that could be achieved through embryo screening for type 2 diabetes: selecting among 10 embryos is expected to reduce absolute disease risk by 12-20% in families where both parents are affected, with similar relative risk reductions across ancestries. These findings establish a framework for implementing PGS in reproductive medicine while demonstrating both the technology’s potential for disease prevention and the methodological standards required for responsible clinical translation.

## Introduction

Disease polygenic scores (PGS) aim to quantify an individual’s genetic liability for developing specific diseases over the course of his or her life ^1^. The development of PGSs relies on genome-wide association studies (GWAS) that quantify the marginal association between primarily common genetic variants and the disease of interest. The collection of large cohorts with genetic and health data has enabled well-powered GWASs^2^, leading to an increasing number of PGSs that attain useful predictive ability across traits and diseases ^3^.

PGSs have shown great promise for risk stratification for common diseases, including breast cancer and coronary artery disease. By 2018, breast cancer PGS identified nearly an order of magnitude more women at equivalent risk as those carrying classic monogenic risk alleles such as those in the BRCA1/2 genes ^4^. Likewise, a coronary artery disease PGS—leveraging GWAS data from biologically related traits such as LDL—classified 20.0% of the population at 3-fold risk compared to the average ^5^. Such results demonstrate the potential for PGSs to be used alongside conventional disease risk factors in risk stratification, with increasing awareness of their utility in informing clinical practice and preventative medicine ^6, 7, 8^.

However, most GWASs to date have relied heavily on European populations^9^. As a consequence, PGS often exhibit substantially reduced predictive performance when applied to genetically distant populations ^10, 11^. Although the underlying causal genetic architecture of traits appears to be broadly conserved across ancestries ^12^, this reduced portability poses both methodological challenges and bioethical concerns regarding the potential exacerbation of existing health disparities ^13^. Addressing this issue requires diversifying study cohorts ^14^ and implementing improved statistical methods for cross-ancestry prediction ^15^.

Although the target of disease GWASs is estimation of direct genetic effects—effects of alleles in an individual on that individual, relevant for within-family prediction—the associations also include contributions from factors not relevant to within-family prediction: indirect genetic effects through the family environment (“genetic nurture” ^16^) and confounding from population stratification and assortative mating ^17^. These factors can lead to attenuation of PGS prediction ability within-families. Although behavioral and social science genetics increasingly quantify and address these biases through within-family designs ^18, 19^, medical genetics typically neglects such controls ^20^, presuming their irrelevance to biologically proximal traits. Consequently, disease PGS are rarely validated within families, with existing studies either focusing primarily on non-disease traits ^19^ or inadequately measuring within-family attenuation ^21, 22^. Furthermore, recent large-scale family-GWAS have revealed imperfect correlations between direct genetic effects for biomedical and disease traits compared to standard population effects (as estimated by non-family-based GWAS), challenging prevailing assumptions^23^.

This is of particular importance for the application of PGS in the context of polygenic embryo screening (preimplantation genetic testing for polygenic traits, PGT-P), which uses genome-wide data to predict embryos’ genetic risk for complex diseases, and which has garnered attention due to its potential to substantially reduce lifetime disease burden ^24^. The disease risk reduction in PGT-P is primarily determined by the direct effect of a PGS (i.e., its association with the trait or disease within-family ^25^). Without stringent validation, including estimation of the PGS direct effect within-family, no trustworthy statement on the utility of a PGS for embryo screening can be made.

Despite these methodological concerns, comprehensive validations of state-of-the-art disease PGS remain scarce. Existing studies seldom systematically benchmark PGS across multiple ancestries, and none employ within-family validation strategies to distinguish direct genetic effects from population-level confounding. To address these gaps, we constructed seventeen disease PGS using state-of-the-art methods aiming at maximising prediction ability, and we rigorously assessed their predictive performance in both population-based and within-family contexts. Additionally, we quantify the attenuation of predictive accuracy along the genetic ancestry continuum. Using consistent metrics, we benchmark our newly developed PGS against existing state-of-the-art academic and commercial scores, finding that our models achieve either superior or statistically equivalent performance. Finally, leveraging the validated within-family predictive ability, we illustrate the practical implications of our approach by demonstrating the potential risk reduction achievable through embryo screening using our type 2 diabetes (T2D) PGS.

## Results

### PGS performance between and within families with genetically inferred European ancestries

We constructed a set of seventeen disease PGSs using a curated set of GWAS summary statistics generated by meta-analyzing individual GWAS studies from FinnGen^26^, the Million Veterans Program (MVP) ^27^, the Global Biobank Meta-analysis Initiative (GBMI) ^28^, trait-specific consortia and the UK Biobank (UKBB) ^29^. For UKBB in particular, we ran GWAS on a subset of self-reported white British individuals with genetically inferred Northwestern European ancestry (hereafter the ‘United Kingdom’ group), holding out related individuals and small test sets for later use in PGS validation. We used SBayesRC ^30^, a method that uses genomic annotations to improve effect estimates, to construct our PGSs.

Performance was initially evaluated using a held-out sample of UKBB individuals genetically inferred to have predominantly European ancestries, using relevant disease outcomes ascertained via electronic health records and self reported disease status (**Supplementary Table 1**). For these validations and for later comparisons, predictive performance was evaluated as liability *R*^2^, or the proportion of variance explained on an underlying, continuous measure of disease predisposition. This quantity relies on the liability threshold model of disease, whereby those with liability exceeding that corresponding to a given population prevalence are considered cases. Among the evaluated PGSs, the highest predictive performance was observed for prostate cancer, type 2 diabetes (T2D), hypertension, and Alzheimer’s disease (AD) (**Figure 1A**, **Supplementary Table 2**). The superior performance of these PGSs can be attributed to distinct factors. T2D and hypertension, which are highly polygenic traits, had the largest effective GWAS sample sizes in our analysis (**Table 1**). Complex diseases demonstrate extensive polygenicity ^31, 32, 33^, whereas cancers, while still polygenic, typically show lower degrees of polygenicity ^34^. Prostate cancer demonstrated the highest predictive accuracy among malignancies, corresponding to its larger GWAS sample size relative to other cancers (**Table 1**). Similarly, the exceptional performance of the AD PGS is likely attributable to the presence of the APOE *ε*4 haplotype with an unusually large effect size for complex diseases, which confers a 2–3 fold increased risk with one copy and 10–15 fold increased risk with two copies ^35^. These findings suggest that both GWAS sample size and the genetic architecture of traits are primary determinants of PGS predictive performance ^36^.

**Figure 1:**
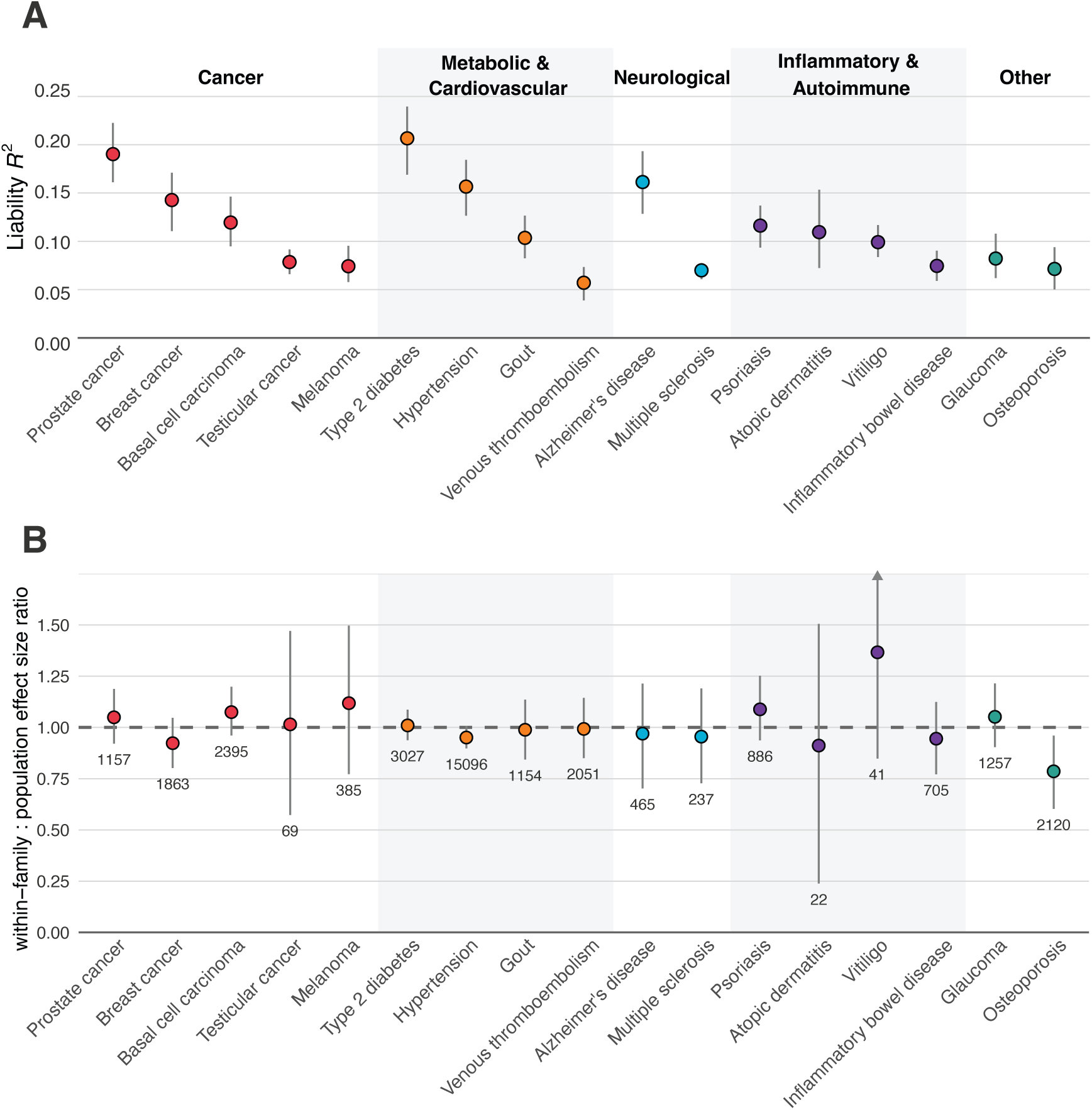
Fraction of the variance in disease liability explained by PGSs between and within families in UK Biobank. A) Variance explained in disease liability by its corresponding PGS on the liability scale. B) The ratio of effect sizes observed in population and within-family regression analyses in identical samples, where the dotted line at 1 indicates identical strength of association between and within families. The numbers below the point estimates indicate the number of probands identified in UKBB as cases. Bands indicate bootstrapped 95% confidence intervals.

**Table 1:**
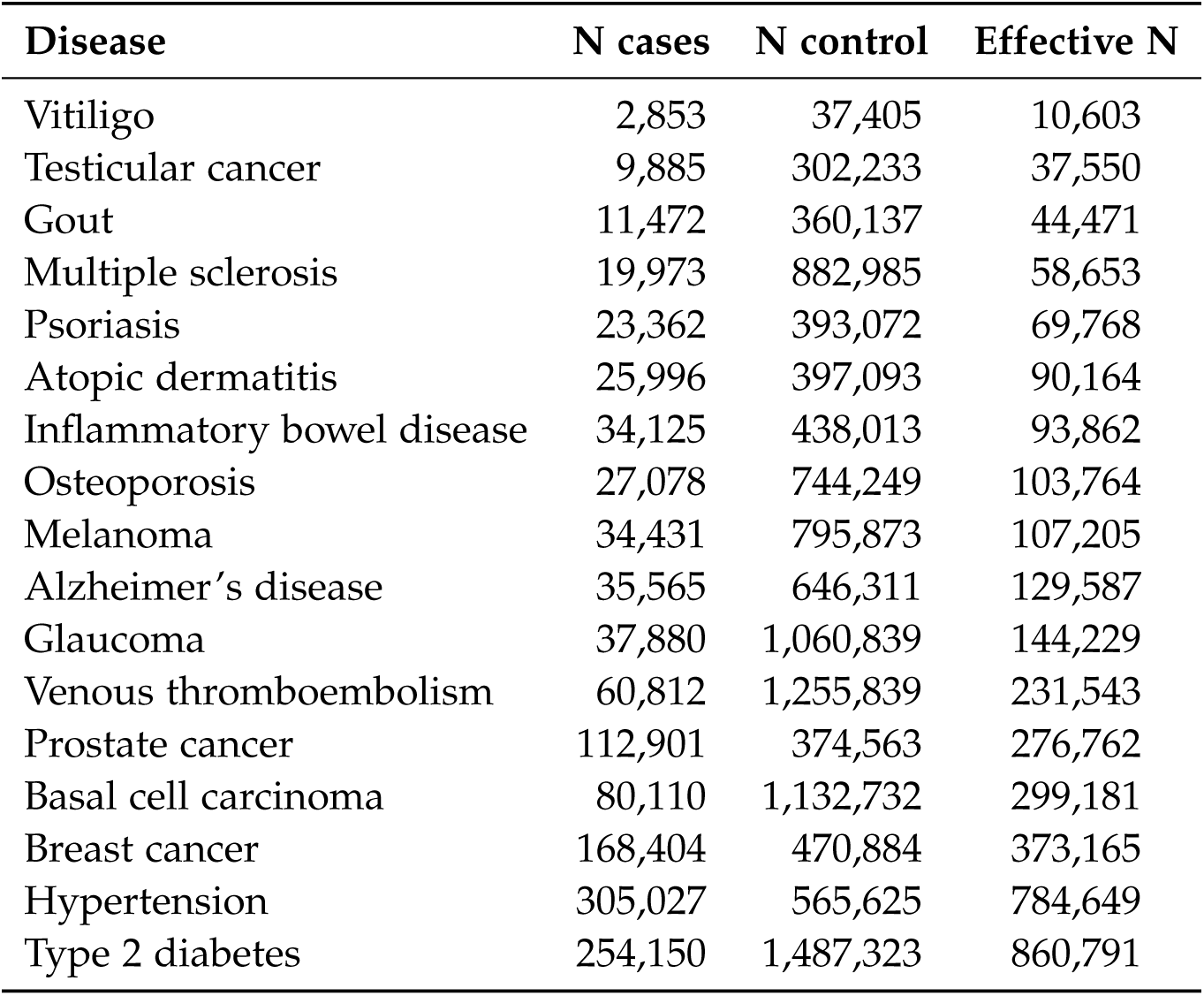
Effective sample size and composition of the GWAS data used in PGS training. Effective sample size was computed as the sum of per-study effective sample sizes following eq. (6) in Grotzinger et al. ^37^ Where input GWASs were themselves meta-analyses, cohort-level information was extracted to compute overall meta-analytic effective sample sizes. If such information was not available, recommendations from Tucker-Drob 2025^38^ were instead implemented.

To estimate direct genetic effects and evaluate the predictive validity of PGSs in an embryo screening context, we performed within-family analyses that control for parental genotypes. We leveraged first-degree relatives in the UK Biobank cohort to impute parental genotypes using the *snipar* package ^18^. This approach enabled us to calculate PGSs for both focal individuals and their (imputed) parents, allowing us to evaluate the degree to which the previously observed effects were driven by direct genetic effects versus other factors.

We found that the within-family predictive power of the PGSs was not significantly different from the population-based associations for sixteen out of the seventeen PGSs considered, with only the PGS for osteoporosis having a nominally significant lower within-family effect (Figure 1B). For the remaining PGSs, all point estimates of the ratio of within-family to population-based effect size estimates were greater than 0.9 and not significantly different from one (*P >* 0.05), suggesting that the predictive ability of these PGSs derives almost entirely from direct genetic effects (**Supplementary Table 6**).

### Attenuation of PGS performance with increasing genetic distance from training cohorts

The reduced performance of PGSs in individuals with ancestries genetically distant from those used in the training GWAS is widely appreciated. This phenomenon is likely due to differences in the linkage disequilibrium structure of genomes with ancestry, with differences increasing with genetic distance, as well as other potential factors ^10^. As the majority of GWASs conducted thus far have used samples of individuals with predominantly European genetic ancestries (including those developed here), it is crucial to evaluate the performance reduction along the continuum of genetic ancestries.

To assess the reduction in PGS performance in samples of individuals with diverse genetic ancestries, we replicated the analyses introduced in Prive et al. ^10^ Firstly, we used an identical PGS generation pipeline as that used for the disease PGSs to generate around 40 to 80 PGSs— with the number varying depending on the sample size for each ancestry — for a variety of additional traits, including continuous biomarker traits for which PGS performance is more easily estimated (**Supplementary Table 1**). Secondly, we constructed a diverse set of eight discretized groups based on genetically-inferred ancestries, using reference panels of individuals with self-reported countries of origin (or religious affiliation in the case of Ashkenazi Jewish individuals). Specifically, we identified groups that genetically resembled self-reported individuals from Poland, Italy, Iran, India, China, the African Caribbean population and Nigeria and those identifying as being of Ashkenazi Jewish descent. Thirdly, we calculated the *R*^2^ for each PGS (e.g., BMI, apolipoprotein A) in the UK Biobank United Kingdom ancestry group and compared it to the *R*^2^ attained in each group. Finally, we regressed the *R*^2^ attained in each group on the value attained in the UK Biobank United Kingdom ancestry group which allowed us to assess the performance reduction factor (Figure 2A).

**Figure 2:**
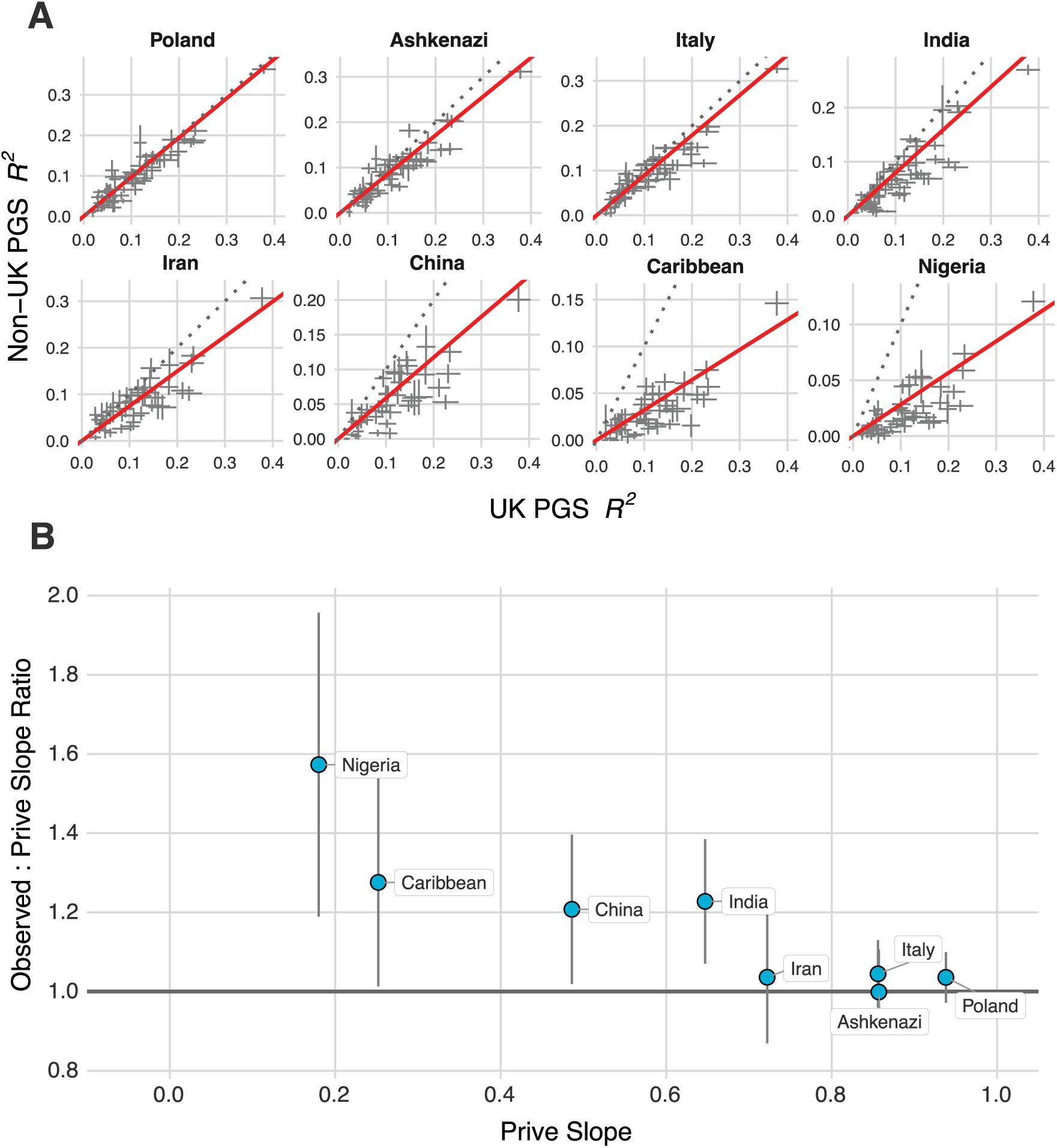
PGS effect size attenuation across ancestries. A) Scatterplots showing the R^2^ attained by a given PGS in the United Kingdom ancestry subsample on the x axis versus that attained in the given non-UK ancestry sample on the y axis. Intervals indicate standard errors, the red lines indicate the Deming regression slope (intercept fixed at 0), and the dotted gray line indicates the y=x line. See **Table 2** for slope values and 95% confidence intervals. B) A scatterplot comparing the effect reduction slopes inferred from Prive et al. ^10^ versus the ratio of the slopes we observed in (A) to Prive et al. for the different groups analyzed. The intervals indicate standard errors for the Deming regression coefficients.

**Table 2:** Attenuation in PGS performance across ancestries. Observed Deming regression slopes with 95 % confidence intervals for the regressions reported in Figure 2 and corresponding slopes from Prive et al.

| Group | Deming regression slope (CI) | Prive et al. slope |
| --- | --- | --- |
| Poland | 0.971 (0.912, 1.03) | 0.938 |
| Italy | 0.894 (0.821, 0.967) | 0.856 |
| India | 0.794 (0.692, 0.896) | 0.647 |
| Iran | 0.748 (0.627, 0.869) | 0.722 |
| Ashkenazi | 0.856 (0.764, 0.948) | 0.857 |
| China | 0.587 (0.495, 0.679) | 0.486 |
| Caribbean | 0.321 (0.255, 0.388) | 0.252 |
| Nigeria | 0.283 (0.214, 0.352) | 0.180 |

We found that PGS performance decreased as the centroid of a given group in common variant PC space increased in distance from the centroid of the training GWAS data (Figure 2). Notably, we found that the relative attenuation in our PGS construction pipeline was less severe than that observed in Prive et al., observing significantly better performance particularly in the more genetically distant groups from the ancestries represented in the training GWASs (Figure 2B). This improvement in portability was likely due to the fact SBayesRC leverages genomic functional annotations that are independent of ancestry, enabling fine-mapping of causal loci, as the authors of SBayesRC similarly observed, albeit on a more limited set of traits ^30^.

### Performance of PGS compared to academic and commercial entities

We next sought to compare the performance of the disease PGS we developed here to those developed by various academic groups and other commercial providers. Thompson et al. (2024) ^39^ and Mars et al. (2022) ^40^ generated several disease PGS that overlap with those generated here and reported the odds ratio associated with a one standard deviation increase in the PGS in UKBB and Finngen, respectively ^26, 29^. Across all scores where we could identify a comparable PGS-disease pair, we found that our scores outperformed or performed as well as those generated by Thompson et al. and Mars et al. (Figure 3A). We note substantially improved performance with respect to breast cancer, melanoma, gout, and multiple sclerosis, with our scores having higher point estimates across all disease with the exception of glaucoma, for which Thompson et al. reported a slightly higher though statistically indistinguishable odds ratio point estimate. Our liability *R*^2^ was on average 28% and 87% higher than those of Thompson et al. and Mars et al., respectively. When compared to the liability *R*^2^ reported by two commercial entities (assuming identical population prevalence), we similarly found the PGSs reported here showed substantially better performance: 122% better than Orchid and 193% better than Genomic Prediction on average (Figure 3B). In several cases, the point estimates suggest the PGSs we trained explain more than two times as much variance in disease liability as explained by the PGSs developed by other commercial entities. Notably, Orchid provided PGS validations for only five out of the 17 traits included here, and we were unable to determine appropriate confidence intervals for their reported performance metrics. When the documented validation methodology of a commercial entity was illegitimate or incompatible with ours, we omitted the score in question (e.g., validation results reported by Orchid for Alzheimer’s disease are therefore not shown as the reported validation includes age as a covariate, which likely had an outsized contribution to their reported performance.) ^41^. While we made extensive efforts to evaluate Nucleus Genomics, we were unable to reconcile their performance estimates with established theoretical and empirical boundaries of polygenic prediction, necessitating their exclusion from our primary analysis (see **Supplementary Note** for technical details)

**Figure 3:**
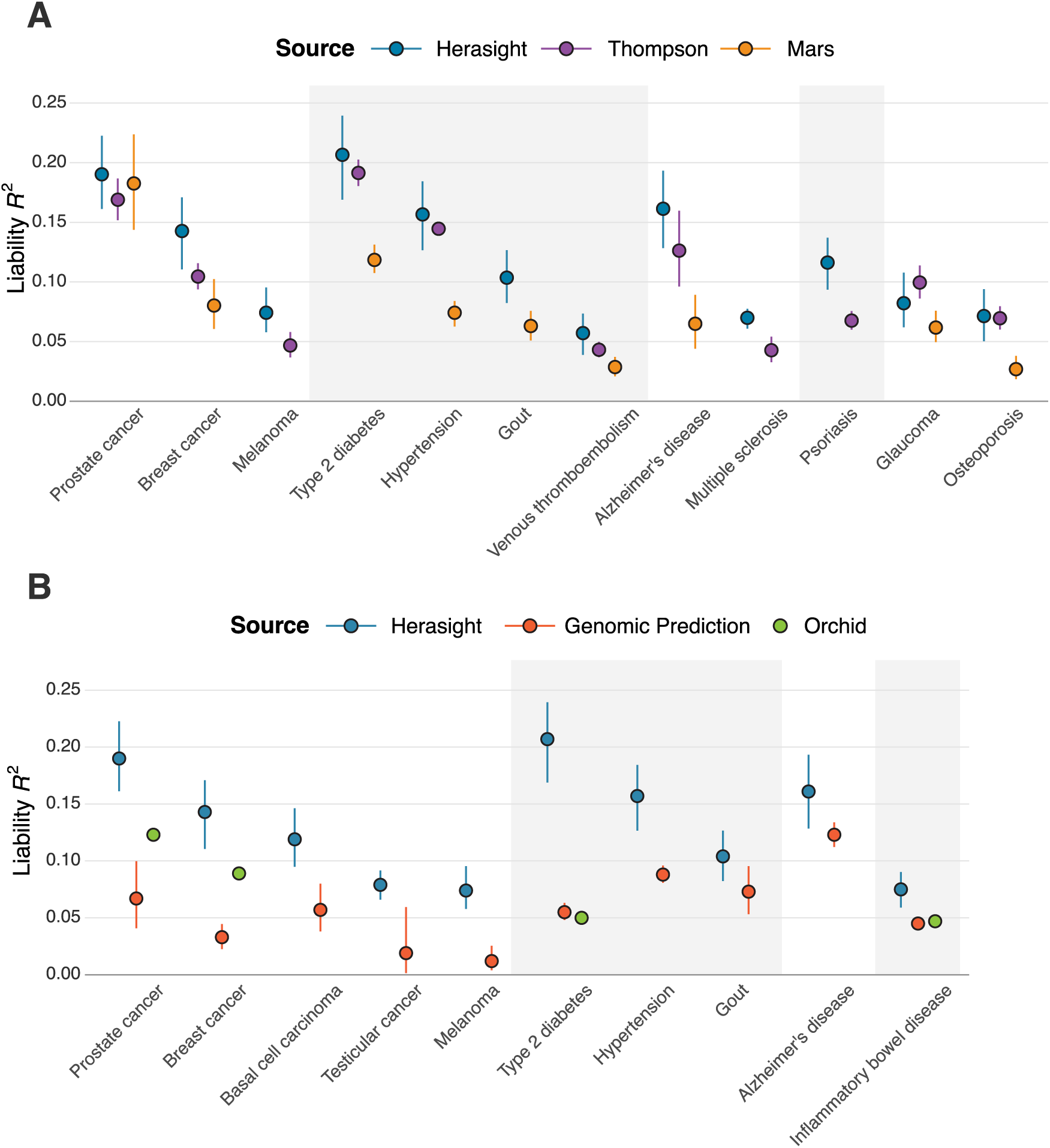
Comparison of PGS performance to external academic and commercial entities. Variance explained in disease liability by its corresponding PGS on the liability scale, comparing results for PGS constructed in the current work by Herasight to those reported by A) two academic groups: Thompson et al. and Mars et al. and B) two commercial entities: Genomic Prediction and Orchid. Bands indicate bootstrapped 95% confidence intervals. Note that we were not able to ascertain 95% confidence intervals for Orchid. Equivalent population prevalences were used in the conversion to liability R^2^ (see **Methods**).

### Case Study: The utility of polygenic embryo screening in reducing lifetime risk of T2D

To demonstrate the utility of our PGSs in the context of embryo screening, we used T2D polygenic screening as a case study. We calculated the expected absolute and relative risk reduction for type 2 diabetes in couples with varied continental genetic ancestries and disease status. These are calculated using mean expected attenuations based on centroids calculated in PC space. Since the true attenuation for any given couple will doubtlessly vary, these calculations are largely illustrative. Even with as few as five embryos, the expected reduction in absolute risk for type 2 diabetes ranges from 5% to 15% depending on parental ancestries and disease status (Figure 4). If screening were applied to twenty embryos, the expected reduction in disease risk becomes substantial to the degree that when both parents are affected, the expected relative risk reduction ranges from 23% to 51% and absolute risk reduction ranges from 14% to 24% depending on parental ancestries.

**Figure 4:**
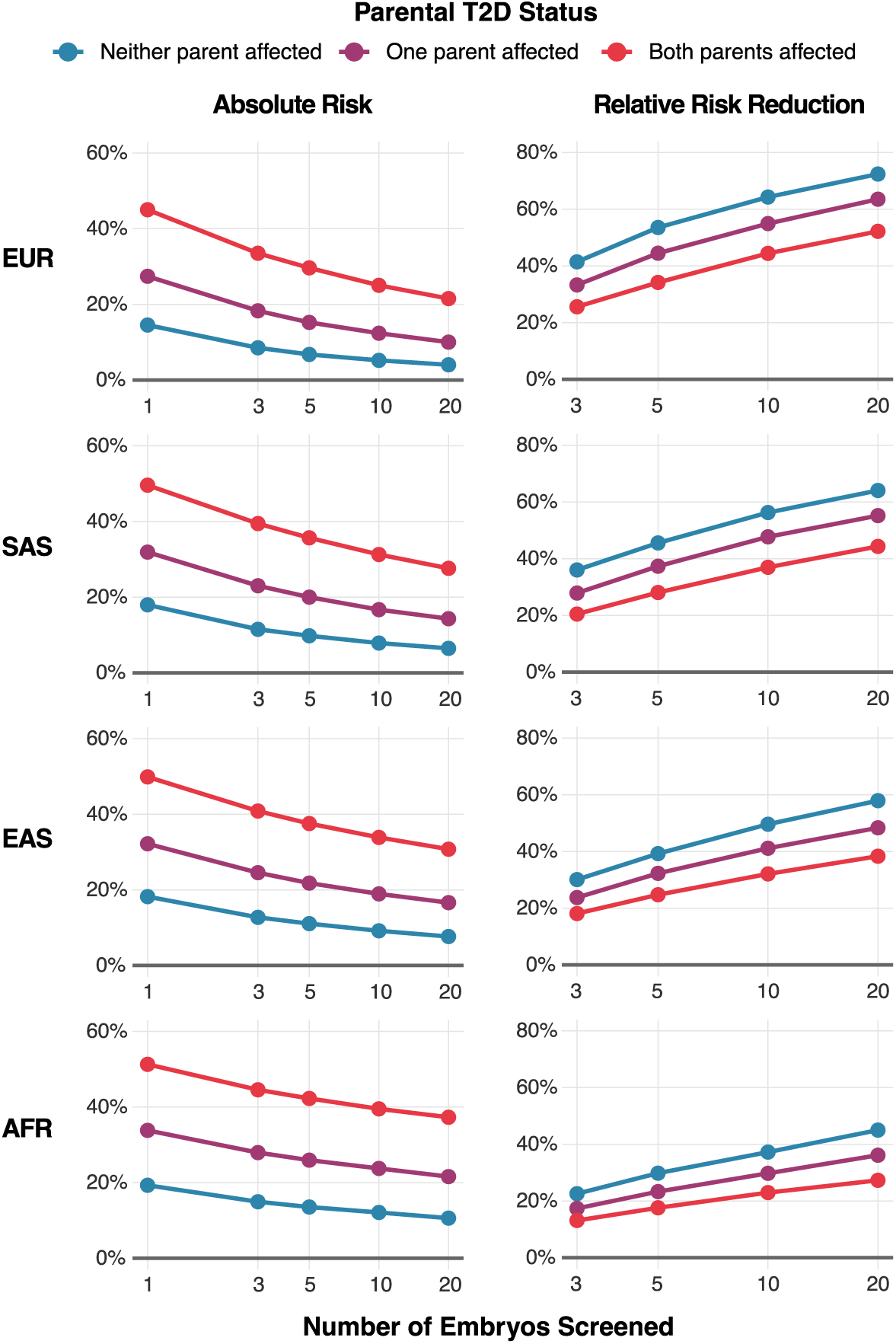
Expected reduction in T2D risk with embryo screening using the Herasight PGS. We calculated the absolute lifetime risk (left) and relative risk reduction (right) parents with different continental ancestries could expect via embryo screening, under various scenarios of parental T2D affectation status and the number of screened embryos (see **Methods**). Lifetime T2D risk estimates are reported in **Supplementary Tables 3** and 4. EUR: European, SAS: South Asian, EAS: East Asian, AFR: African genetic ancestries.

Another determinant of potential risk reductions is the predictive power of the PGS, which as noted above varies both by target disease and model developer. Table 3 displays absolute and relative risk reductions for a hypothetical couple of European continental genetic ancestry in which either the father or mother is affected. Selection among five embryos in this scenario yields relative risk reductions exceeding 40% where PGS liability *R*^2^ is high, but commensurately lower reductions of 21% to 36% where the PGS only captures a few percentage points of liability variance.

**Table 3:** Expected reduction in disease risk with embryo screening using validated PGS from providers of preimplantation genetic testing for polygenic diseases (PGT-P) (Herasight, Orchid, and Genomic Prediction (GP)). We calculated the absolute and relative risk reductions a couple of European continental genetic ancestry with one parent affected could expect when screening five embryos using various PGS models. Lifetime breast cancer, prostate cancer and T2D risk estimates are reported in **Supplementary Table 3**. “Population baseline” refers to the risk pertaining to all families in the population and “Family baseline” only to those with one parent affected by the disease, which are therefore higher than the former for each disease.

| Disease | Company | PGS<br>$R^2$ (%) | Population<br>baseline (%) | Family<br>baseline (%) | Absolute risk<br>reduction (%) | Relative risk<br>reduction (%) |
| --- | --- | --- | --- | --- | --- | --- |
| Breast cancer | Herasight | 14.3 | 13.9 | 19.8 | 8.1 | 41.0 |
|  | Orchid | 8.8 | 13.9 | 19.8 | 6.4 | 32.3 |
|  | GP | 3.3 | 13.9 | 19.8 | 4.1 | 20.6 |
| Prostate cancer | Herasight | 19.0 | 12.6 | 24.5 | 10.8 | 44.2 |
|  | Orchid | 12.3 | 12.6 | 24.5 | 8.8 | 36.2 |
|  | GP | 6.7 | 12.6 | 24.5 | 6.7 | 27.4 |
| Type 2 diabetes | Herasight | 20.7 | 21.4 | 27.5 | 12.1 | 43.8 |
|  | Orchid | 5.0 | 21.4 | 27.5 | 6.1 | 22.0 |
|  | GP | 5.5 | 21.4 | 27.5 | 6.6 | 23.8 |

## Conclusion

We have generated PGSs for seventeen diseases spanning several disease groups. We demonstrated state-of-the-art performance relative to academic and commercial benchmarks across diseases, attaining superior or statistically indistinguishable performance to other groups in all cases. Our work represents the first systematic within-family validation across multiple disease PGS at this scale, demonstrating that predictive accuracy is predominantly driven by direct genetic effects for sixteen of seventeen diseases evaluated. The exception, osteoporosis, serves as a proof of principle that PGS prediction of disease traits can exhibit reduced performance within-family due to confounding, underscoring the importance of within-family validation for any PGS intended for embryo screening. A PGS exhibiting reduced performance within-family can still be used for embryo screening, but the reduced prediction ability needs to be accounted for when calibrating predictions By applying Bayesian mixture models to more than seven million SNPs while utilising genomic functional annotations ^30^, we observed improved cross-ancestry portability compared to previous approaches, with predictive accuracy maintained at higher levels in individuals genetically distant from the training data. Although this represents progress toward addressing disparities in PGS performance, achieving more equal accuracy across all ancestries remains a central challenge requiring continued development of diverse cohorts and methodology.

Our results show that polygenic embryo screening holds promise for reducing disease risk, particularly when a family history of disease is present. As the T2D case study illustrates (Figure 4), the absolute risk reductions are most pronounced when one or both parents are affected. However, current PGS implementations, including ours, primarily capture common variant contributions to disease risk. Family history often additionally reflects the segregation of rare, high-effect variants that substantially influence disease liability but are not captured by PGSs. Comprehensive risk assessment in the embryo screening context will require integrated models that combine polygenic background from common variants, high-effect rare variants (particularly critical in families with strong disease history), and explicit family history information. The preimplantation genetic testing (PGT) setting presents unique methodological challenges for such integration, including constraints on DNA quantity from embryo biopsies and the need for accurate imputation and variant calling across the allele frequency spectrum. Future work should prioritize improving the calling of all classes of genetic variation in embryos, ultimately enabling more accurate risk stratification that reflects the full genetic architecture underlying disease risk. Our findings suggest several directions for advancing the field. The consistent within-family performance across most diseases indicates that continued investment in larger, more diverse GWAS should yield direct benefits. The improved portability achieved through methodological enhancements demonstrates that technical innovation can help address existing disparities even with current data limitations. The validation approach presented here, encompassing population-based performance, within-family analyses, ancestry portability assessment, and systematic benchmarking, may serve as a useful framework as the field moves toward clinical implementation.

Important challenges remain in characterizing pleiotropic effects, integrating rare variant information, and ensuring sufficient performance across ancestries. Similarly, the probabilistic nature of polygenic risk requires careful communication strategies that contextualize individual PGS-predicted disease risk that appropriately considers relevant family history to produce calibrated absolute risk estimates, which our analyses suggest are inadequately executed by other commercial providers (see **Supplementary Note**).

Despite these challenges, our results suggest that polygenic embryo screening, when implemented with appropriate validation, offers meaningful potential for reducing disease risk in future generations. Continued methodological development, coupled with open reporting and rigorous validation, will be essential for realizing this potential while ensuring broad access.

## Methods

### UK Biobank

The UK Biobank (UKBB) is a large population-based study consisting of nearly 500,000 individuals with genotypes and linked health records recruited from 22 sites throughout the UK ^14^. The array-based version of the dataset imputed by Wellcome Trust Centre for Human Genetics (WTCHG; UKBB field 22828) was used during the first phase of the present study consisting of the development of the PGS models. To ensure quality control of this version of the UKBB genotypic and phenotypic data, we excluded individuals with outlier heterozygosity (UKBB field 22027), ten or more third-degree relatives also present in the data and a genotype missingness rate greater than 3%. To retain as much trait-associated variation as possible while minimizing artifacts, genetic variants were excluded only when either Hardy-Weinberg (HWE) *p*-value or MAF was less than 1 × 10^−50^ and 1%, respectively. The final set of included variants for use in UKBB GWAS consisted of 6,667,399 biallelic SNPs overlapping with those comprising the SBayesRC variant panel ^30^. We used the whole-genome sequence-based version of the UKBB dataset for PGS model validation during the second phase of the present study. Samples retained after quality control of the imputed version of the dataset were likewise retained for use in this second phase. Minimal filtering of variants passing GraphTyper’s FILTER field with AAScore *>* .8 and MAF *>* .1% resulted in a final set of 7,153,533 biallelic SNPs again overlapping with those comprising the SBayesRC variant panel.

### UKBB ancestry inference

Ancestry of UKBB samples passing above sample and genotype filters was inferred using the country-of-origin method described in Prive et al. ^10^. Specifically, we accessed pre-computed principal component (PC) centroids for the subsets of UKBB participants hailing from the Caribbean, China, India, Iran, Italy, Poland, Nigeria and the United Kingdom as well as that for self-reported Ashkenazi Jews. We then matched ancestries to these centroids by computing the Euclidean distance between the first 16 PCs of QC-passing samples and the seven ancestry centroids; samples were assigned to the country-of-origin group of the nearest centroid. Samples matched to the “United Kingdom” ancestry group were further filtered to those individuals also reporting “Caucasian” genetic grouping (UKBB field 22006) and “British” ethnic background (UKBB field 21000).

### GWAS summary statistics and meta-analysis

To attain the largest possible discovery sample sizes for PGS training, we collated GWAS summary statistics from several classes of sources: recently published GWAS, trait-specific genomics consortia, GBMI ^28^, FinnGen ^26^, healthcare registry GWAS, and internally-run UKBB GWAS. In cases where summary statistics were not immediately available, corresponding authors were contacted for provision of UKBB-left-out data and were acknowledged where re-analysis was required.

All summary statistics files were first harmonized to GRCh37 and corrected for errors in per-SNP sample size ^42^ following the recommendations from Tucker-Drob (2025) ^38^, using cohort-level information where possible. Corrected summary statistics files were then meta-analyzed with inverse variance weighting in METAL ^43^ where LDSC-estimated ^44^ genetic correlations between the available sources exceeded ∼ 0.8. Where possible, UKBB GWAS were run with either REGENIE ^45^ or fastGWA^46^ on a discovery sample consisting only of United Kingdom ancestry samples not overlapping with pre-selected test sets.

### Construction of PGS

Internal PGS were built by providing our meta-analytic summary statistics as input to SBayesRC ^30^, a recently published Bayesian method leveraging information from functional genomic annotations. Training consisted of two stages: a first stage to perform basic quality control on input association statistics and to impute associations from untested SNPs as well as a second stage to perform Markov Chain Monte Carlo parameter estimation. Resulting scoring files included weights from the ∼ 7.3M SNPs comprising the SBayesRC variant set provided by the authors of the method.

### Literature-derived lifetime risk estimates

Lifetime risk estimates were estimated using available measures of lifetime risk, lifetime prevalence, and cumulative incidence. Data were collected from the National Cancer Institute’s Surveillance, Epidemiology, and End Results (SEER) Program ^47^, Global Burden of Disease Study ^48^, CDC ^49^, and various other sources (see **Supplementary Tables 3** and **4**). In many cases multiple measures of lifetime risk were averaged. Population prevalence estimates were taken as the average of the male and female prevalence estimates for conditions that present in both sexes.

### PGS validation and conversion of observed to liability ***R*^2^**

Polygenic scores (PGS) were evaluated in **R** with the lm() function from the **R** *stats* package. For continuous traits we fit ordinary least-squares models in 1,000 unrelated UK Biobank participants of “United Kingdom” genetic ancestry, adjusting for age and the first ten PCs. For diseases with sufficient numbers of cases to permit test-set exclusion and an internal UKBB GWAS, PGS were validated in test sets consisting of 600 UKBB cases and 1,000 UKBB controls using a linear probability model (lm() on a 0/1 outcome) with PGS residualized on age and the first ten PCs. The estimated observed *R*^2^ from these models was then converted to liability *R*^2^ using the transformation of Lee et al. ^50^ (“*Observed-scale R*^2^ → *liability-scale R*^2^” in **Supplementary Table 5**). Standard errors were bootstrapped with 2,000 replicates.

### Within-family PGS validation

Additional PGS validations were performed in the subset of UKBB participants for whom genotypes of at least one other parent or sibling were also available to permit Mendelian imputation of parental genotypes using the *snipar* package ^18^. Such samples were identified using KING ^51^ and filtering to non-monozygotic relatives belonging to the United Kingdom ancestry group. In total 40,943 probands were included in the analysis. Variants were further filtered to those amenable to *snipar*-based genotype imputation with INFO *>* 0.99, resulting in a reduced set of 4,065,217 biallelic SBayesRC SNPs for within-family PGS scoring.

Binary outcomes were analysed with logistic regression, fitted in **R** via the *lme4* package glmer() function using the binomial probit link and modeling phenotypic correlations between siblings by fitting family-wise random intercepts. A first “population” model regressed the phenotype on age, sex, the third and fourth principal components as well as offspring PGS; a second “within-family” specification added the maternal and paternal snipar-imputed parental PGSs to separately estimate the non-transmitted component following the model introduced by Young et al. (2022, Equation 1) ^18^ (estimated population effects, direct effects, average NTCs and spousal PGS correlations are reported in **Supplementary Table 6**). To estimate the attenuation of PGS performance within-family, the ratio of the offspring PGS coefficients from the within-family and the population models was calculated for each trait and its standard error approximated via family-wise bootstrap with 2000 replicates.

### Relative performance estimation

Relative performances were computed as the slope of a Deming regression of estimated variance explained in each non-“United Kingdom” group on that of the “United Kingdom” group for each trait. Regressions consisted of around 40 to 80 pairs of PGS performance estimates for the focal traits and diseases listed in **Supplementary Table 1**.

### Comparison of PGS performances against those of academic PGS developers

To benchmark our PGS validation results against those previously reported in the academic literature, we extracted and converted performances recently published in Thompson et al. and Mars et al., which develop and validate PGS in UKBB and FinnGen, respectively. Performances from the former are provided in the publication’s supplementary Table S3: Performance (disease) and in Table S4 of the latter. Both sources reported performances as odds ratios per standard deviation of PGS, which were converted to liability *R*^2^ using known conversions (i.e., by sequentially applying the “*Odds-ratio per 1 SD of PGS (OR)* → *Cohen’s d*”, “*Cohen’s d* → *observed-scale R*^2^” and “*Observed-scale R*^2^ → *liability-scale R*^2^” conversions shown in **Supplementary Table 5** using the trait-specific lifetime risks and sample prevalences given in **Supplementary Tables 3**).

### Comparison of PGS performances against those of commercial PGS for PGT-P providers

We likewise benchmarked our results against those previously claimed by commercial providers of PGS. Given a growing emphasis on the use of PGS for PGT-P, we extracted performances published online in whitepapers by Orchid Health ^52^ and by Genomic Prediction (now Lifeview) as Widen et al. ^21^, acknowledging that Genomic Prediction has since displayed risk reductions for a new PGS set whose validation performances have not been made public ^53^. Both sources reported performances as AUROC, but only the latter computed parameter uncertainties. Reported values were converted to liability *R*^2^ using known conversions (i.e., by sequentially applying the “*Area under ROC curve (AUC)* → *Cohen’s d*”, “*Cohen’s d* → *observed-scale R*^2^” and “*Observed-scale R*^2^ → *liability-scale R*^2^” conversions shown in **Supplementary Table 5** using the trait-specific lifetime risks and sample prevalences given in **Supplementary Tables 3** and **7**, respectively). For breast cancer, type 2 diabetes and inflammatory bowel disease, Orchid also reported performance as odds ratios per standard deviation of PGS, which was used for conversion to liability *R*^2^ in place of an incomparable AUC metric computed using both PGS and additional covariates. In particular, the inclusion of age as an additional covariate in Orchid’s sole published benchmark for Alzheimer’s disease likely inflated the resulting AUC, precluding meaningful comparison between this and otherwise comparable estimates.

### Simulations of expected gains from use of PGS models in PGT-P

Absolute and relative risk reductions achievable when applying the T2D PGS model in PGT-P were determined via simulation following the family-based variance decomposition first developed and implemented by Lencz et al. ^24^ Specifically, the portion of the genetic component captured by the PGS and shared between embryos was drawn as 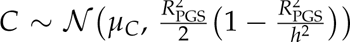 with *R*_PGS_ reduced according to the estimated relative performances corresponding to the parental ancestries (e.g., *R*^2^ reduced from *R*^2^ = (20.7% + .32 × 20.7%)/2 = 13.7% and *R*^2^ = .59 × 20.7% = 12.2% for a EUR/AFR couple and an EAS/EAS couple, respectively). *µ_C_* depended on the average full genetic components of the parents (*g_m_* and *g_f_*) and the trait heritability such that *µ_C_* = *R*^2^ /*h*^2^ × (*g_m_* + *g_f_*)/2. In turn, *g_m_* and *g_f_*were allowed to separately depend on each parent’s disease status, ancestry- and sex-specific lifetime risk of disease and the disease heritability. These were obtained via accept–reject sampling (ratio-of-uniforms implementation). The portion of the genetic component captured by the PGS and unshared between embryos was accordingly drawn as *X* ∼ *N*(0, *R*^2^ /2) and the full PGS for each embryo computed as pgs = *x* + *c*. The uncaptured, unshared genetic component *G*_res_ ∼ *N*(0, (*h*^2^ − *R*^2^)/2) and the environmental residual *E*_res_ ∼ *N*(0, 1 − *h*^2^) were then added to the full average parental genetic component, the unshared, captured genetic component and the environmental residual to compute the corresponding disease liability for each embryo (i.e., *l* = (*g_m_* + *g_f_*)/2 + *g*_res_ + *x* + *e*). Embryos were designated as disease cases where liability exceeded the disease threshold *T* = Φ^−1^(1 − *K*), which in turn depended on embryos’ ancestry- and sex-specific lifetime risk of disease *K*.

Embryo PGS, liability and disease status were simulated in 150,000 batches of 3, 5, 10 and 20 embryos for each unique combination of parental ancestries, parental disease status (i.e., father and/or mother affected) and batch size (400 unique combinations in total). Simulated prevalence of disease with and without selection was computed as the fraction of diseased embryos with the lowest PGS in each batch (*P*_selection_) and the overall fraction of diseased embryos (*P*_population_), respectively. Absolute risk reductions were then calculated as *P*_population_ − *P*_selection_ and relative risk reductions as (P_population_ − P_selection_)/P_population_.

### Inference of implied liability variance explained from risk predictions

To quantify the implied performance of a PGS from disease risk predictions, we applied the liability threshold model to infer the liability variance explained (liability *R*^2^ or *R*^2^) from combinations of PGS *z*-scores, population prevalence, and predicted individual risk. Under the liability threshold model, disease liability *L* is modeled as *L* = *G* + *E*, where *G* represents the genetic component distributed as *G* ∼ *N*(0, *R*^2^), *E* represents the environmental component distributed as *E* ∼ *N*(0, 1 − *R*^2^), and *G* and *E* are assumed to be uncorrelated.

For a binary disease trait with population prevalence *K*, the liability threshold *T* is defined as *T* = Φ^−1^(1 − *K*), where Φ^−1^ denotes the inverse standard normal cumulative distribution function. The probability of disease for an individual with standardized PGS value *g* is given by:

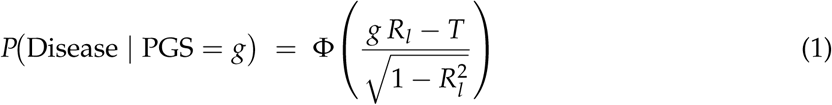

To infer the implied *R*^2^ from reported risk predictions, we solved numerically for the value of *R*^2^ that satisfies:

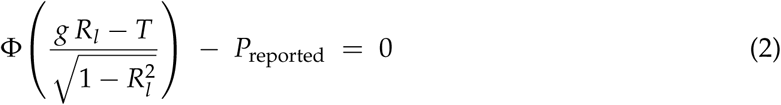

where *P*_reported_ represents the individual disease risk reported for an individual. This equation was solved using bisection root-finding with tolerance 10^−12^, implemented in **R**. This approach enabled us to quantify the predictive performance implicitly claimed by risk predictions made for a given individual.

## Data Availability

The data generated in this study are proprietary and are not publicly available.

## GWAS summary statistics

We thank the following individuals and groups: the Type 2 Diabetes Global Genomics Initiative (T2DGGI) for generation of association summary statistics without UKBB; the International Multiple Sclerosis Genetics Consortium (IMSGC) for provision of GWAS summary statistics; the participants and investigators of the FinnGen study; all other authors of GWAS used in PGS training for upholding data sharing norms in the GWAS field.

## Datasets

This research has been conducted using the UK Biobank Resource under Application Number #103244. This work uses data provided by patients and collected by the NHS as part of their care and support.

## Author contributions

**Spencer Moore:** Conceptualization; Data curation; Formal analysis; Software; Visualization; Writing — original draft; Writing — review & editing

**Ivan Davidson:** Investigation; Writing — review & editing

**Jonathan Anomaly:** Writing — review & editing **Jeremiah H. Li:** Writing — review & editing **Mohammad Ahangari:** Writing — review & editing **Lauren Moissiy:** Writing — review & editing

**Michael Christensen:** Supervision; Project administration; Funding acquisition; Writing — review & editing

**Alexander Strudwick Young:** Methodology; Writing — review & editing

**David Stern:** Conceptualization; Methodology; Visualization; Supervision; Writing — review & editing

**Tobias Wolfram:** Conceptualization; Methodology; Supervision; Project administration; Writing — review & editing

## Supplementary Note: Assessment of Nucleus Genomics’ polygenic score calibration and performance

In the course of conducting a comprehensive benchmark of polygenic embryo screening capabilities across commercial providers (see Figure 3B), we initially sought to include Nucleus Genomics (hereafter referred to as Nucleus), a direct-to-consumer genetic testing company providing disease risk predictions through physician reports, that recently expressed its intent to offer PGT-P. Given the absence of published performance validations by Nucleus, we performed an independent assessment using publicly available documentation and a set of five reports (three from European ancestry and two from non-European ancestry customers), made available to us with customer consent, from up to June 2025, in order to compare their performance to the other commercial providers.

**Supplementary Note Table 1:** Nucleus disease PGSs with low numbers of variants with implied and validated liability *R*^2^ values. *Substantial overlap between the UKBB-containing PGS training sample and UKBB test set will have inflated the estimated liability R^2^.

| Disease PGS | Number of variants | Median implied EUR liability $R^2$ | UKBB validation EUR liability $R^2$ (SE) |
| --- | --- | --- | --- |
| Gastric cancer | 6 | 0.1% | 0.04% (.06) |
| ADHD | 12 | 4.3% | 0.007% (.30) |
| OCD | 12 | 9.5% | 0.004% (.05) |
| PCOS | 14 | 8.1% | 2.0% (.62) |
| Endometriosis | 15 | 8.9% | 0.011% (.10) |
| Anxiety disorders | 20 | 8.3% | 0.21% (.25) |
| Severe acne | 20 | 8.1% | 1.3% (.49) |
| Migraine | 21 | 17.6% | 0.65% (.40) |
| Hypertension | 42 | 11.7% | 0.65% (.46) |
| Parkinson's Disease | 50 | 13.9% | 0.73% (.30) |
| Asthma | 152 | 10.7% | 3.8% (.90) |
| Multiple sclerosis | 283 | 10.2% | 3.3% (.36) |
| Coronary artery disease | 502 | 15.2% | 6.2%* (1.3) |

Our evaluation showed considerable variability in the composition of the polygenic scores (PGS) provided by Nucleus. Nine of their disease PGSs appeared to be open-source models from the PGS catalog, almost all being five or more years old. Other disease predictions relied on variant sets notably smaller than typically required to adequately capture polygenic signals (Supplementary Note Table 1). For example, the polygenic score for attention-deficit hyperactivity disorder (ADHD) reportedly consisted of 12 variants, and a Parkinson’s disease score contained 50 variants. These variant counts fall substantially below conventional benchmarks for using PGSs to predict traits with highly polygenic architectures. Further analysis suggests these smaller sets likely correspond to top-ranking genome-wide significant variants reported in previous genome-wide association studies (GWAS), an approach which typically yields limited predictive accuracy for highly polygenic diseases. Most of the scores seem furthermore to be identical to scores used by Nucleus’ open source precursor Impute.me, implying that updates to their score set since the inception of the project as a for-profit business have been limited (see Supplementary Table 8). The only publicly documented update seems to have occurred on May 21st, 2025, when the company swapped a T2D PGS consisting of 171,249 variants (PGS Catalog ID: PGS000036) for another containing 1,259,754 variants (PGS Catalog ID: PGS002308).

To further assess the calibration and validity of their risk predictions, we converted reported absolute and relative risks into implied liability-scale variance explained (liability *R*^2^) via standard liability-threshold transformations (**Methods**). Across the five customer reports, we noted substantial variability and inconsistencies in the implied predictive performance, with, for example, schizophrenia having implied liability *R*^2^ ranging from 0% to 5% (**Supplementary Table 9**). We also noted several examples where the liability *R*^2^ exceeded realistic expectations given the number of variants included in the PGS. For instance, the hypertension PGS, which included only 42 variants, had an implied median liability *R*^2^ values exceeding 10% (**Supplementary Table 9**). We were unable to identify public documentation provided by Nucleus that would explain these results. Available reports included statements qualifying the predictive performance of PGS models (“*Science’s current ability to use common DNA variants to predict a person’s risk of developing [disease] is limited [or ‘extremely limited’ for some diseases].*”) for gastric cancer, depression, ovarian cancer, osteoarthritis, bipolar disorder and colorectal cancer.

**Supplementary Note Figure 1:**
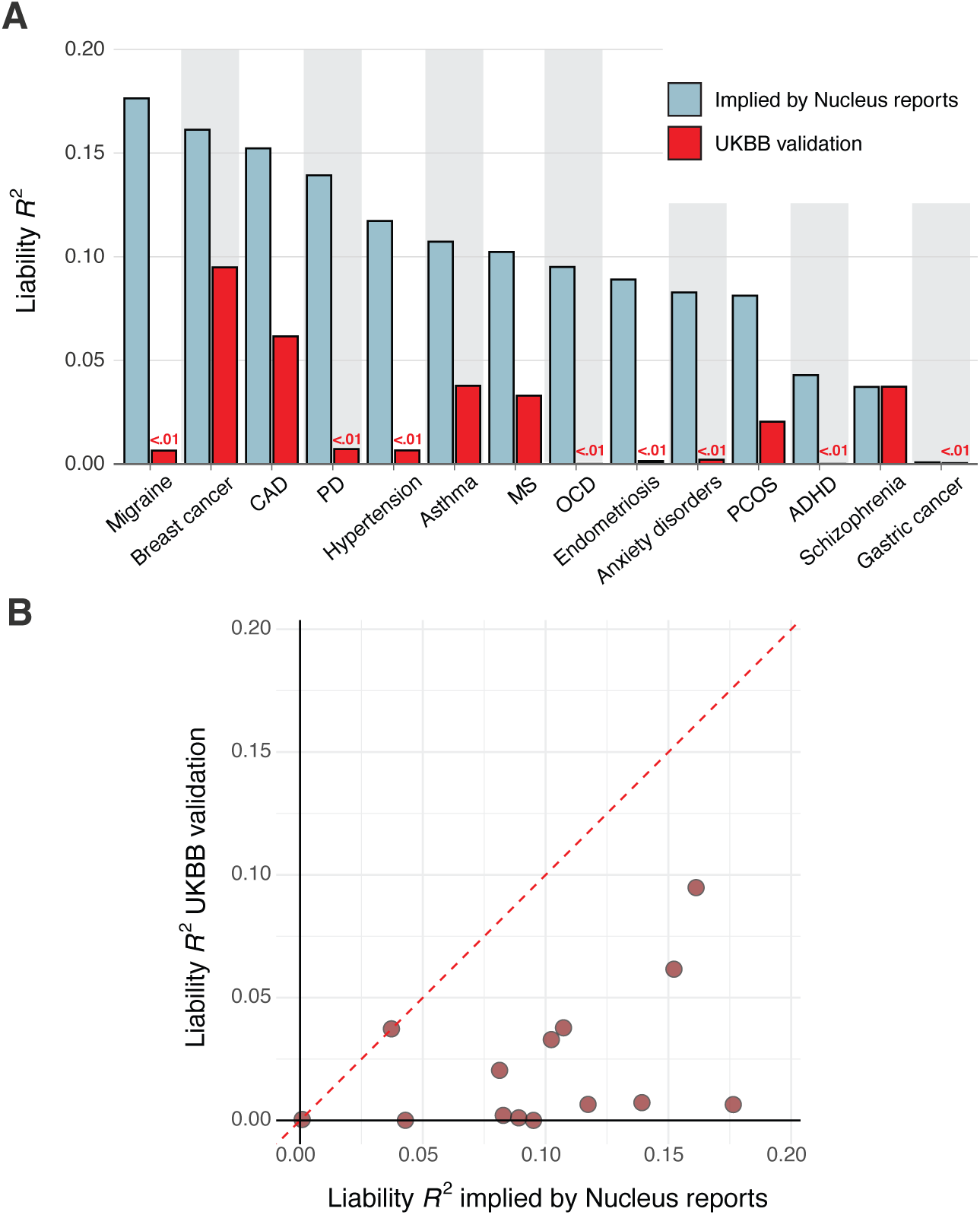
Comparison of Nucleus-implied median liability *R*^2^ (blue) with UKBB validation for the best-matched score (red). (A) Bar chart of median values. (B) Scatterplot of median Nucleus values versus UKBB values; the dashed red line marks the identity *y* = *x*.

We compared Nucleus’ reported risk predictions against our independent UKBB validation for PGS models we could reliably match to their likely scores (see Supplementary Table 8 for the best-matched PGS model source). For nearly all diseases, the median implied performance derived from Nucleus Genomics reports collected from customers of European ancestry was consistently (and often substantially) higher than our independently validated performance estimates (Supplementary Note Figure 1). Notably, for two PGSs where we found literature-based performance estimates, the estimates from our own validation in UKBB were highly concordant (Supplementary Note Table 2). This concordance suggests that our independent validation was consistent with externally conducted validations. Overall, we found that the PGSs likely used by Nucleus display substantially lower predictive performance than that implied by their customer reports.

We also performed a preliminary analysis of Nucleus’ advertised risk reductions when applying their PGS models in PGT-P. To this end, we extracted data from Nucleus’ online risk reduction calculator ^1^, which, for nine diseases, displays the disease’s population prevalence alongside the risk reduced when selecting the embryo with the lowest predicted risk among two to five embryos. The interpretation of these reductions is complicated by the use of prevalences pertaining to various narrow subsets of the population. For example, the reported coronary artery disease (CAD) risks were “calculated for males with one major risk factor,” where major risk factors included smoking, having diabetes, blood pressure (160/100 or above), and high cholesterol (240 and above). The source (Table 3 of Lloyd-Jones et al. ^2^) matches that cited in Nucleus’ physician reports. For other traits, the population prevalences were uncited but identically matched those in the respective physician reports.

**Supplementary Note Table 2:** Comparison of PGS liability *R*^2^ and 95% confidence intervals estimated in our UKBB validation versus those reported in the literature (see Supplementary Table 8 for more details).

| Disease PGS | UKBB validation | Reported in literature |
| --- | --- | --- |
| Breast cancer | 0.095 (.069, .12) | 0.093 (0.087, 0.10) |
| Schizophrenia | 0.037 (.027, .050) | 0.035 (0.015, 0.062) |

This raises the question of whether the reported embryo-level risk reductions apply exclusively to embryos whose future cardiovascular profiles match the described high-risk subset (e.g., males with at least one major risk factor). If so, the interpretation is unclear, as embryo phenotypes cannot be known in advance—and many risk factors themselves partially reflect underlying genetic liability for CAD. If Nucleus derived risk reductions using such narrowly defined subgroups, the reported reductions would not reflect expected risk in a general embryo population. Moreover, if these subsets were chosen based on convenience (e.g., the first figure from the CAD PGS source publication), it raises further questions about methodological rigor.

We collated these prevalences, risk reductions, and Nucleus’ cited disease heritabilities in order to simulate the risk reductions permitted by a given PGS model liability *R*^2^ using the same approach described above (“*Simulations of expected gains from use of PGS models in PGT-P*”). We conducted a limited grid search up to ±10% on either side of the median liability *R*^2^ previously inferred from physician reports, with step sizes of 0.5%, selecting the three estimates best minimizing the sum of reported minus simulated risk reductions across the displayed embryo batch sizes (**Supplementary Table 10**). Although broadly informative, this procedure did not permit confident mapping of embryo-level risk reductions to specific PGS models reconstructed from physician reports.

Nonetheless, in several instances the implied liability *R*^2^ estimates from embryo selection fell within the range inferred from adult reports. For instance, embryo versus adult median European liability *R*^2^ values were 10–11% versus 9.2% for age-related macular degeneration, and 8–9% versus 8.1% for PCOS (Supplementary Table 10). However, substantial discrepancies emerged for traits such as schizophrenia, rheumatoid arthritis, and endometriosis, where inferred embryo-selection liability *R*^2^ values differed considerably from those derived from adult reports. Though such comparisons rely on untestable assumptions concerning Nucleus’ methodology, either possibility—namely that the same questionable PGS models are used in both adult and embryo products or that an unknown set is employed solely for the latter—erodes confidence in the accuracy and reliability of Nucleus’ reported risk reductions. This uncertainty underscores the critical need for calibrated risk estimates, transparent reporting of model validation, and full disclosure of methodology and its assumptions.

Finally, given the above evidence in favor of substantially inflated predictive performances of Nucleus’ PGS models, we sought to informally evaluate hypotheses that might explain the magnitude and breadth of the likely errors. One possible explanation is suggested by the similarity of the implied liability *R*^2^ values to estimates of loosely related parameters in the source GWASs used for PGS training. For example, the median liability *R*^2^ of 17.6% implied by Nucleus’ migraine risk predictions for European customers is closest to a measure of the *SNP heritability* of 14.6% cited in Supplementary Figure 6 of Gormley et al. ^3^, the PGS’s corresponding 2016 migraine GWAS. Crucially, this parameter indexes the proportion of liability variance explained by the additive effects of *all* common causal variants ^4^ rather than just those whose GWAS effects are imprecisely estimated and included in a 21-SNP score. Likewise, Nucleus’ median implied *R*^2^ of 4.3% for ADHD better matches the headline PGS *R*^2^ ∼ 5.5% returned from the leave-one-cohort-out PRS validation in Demontis et al. ^5^; however, this result relates to PGS built with liberal pruning and thresholding, which therefore contain hundreds of thousands of SNPs in addition to the 12 included in Nucleus’ ADHD model. A similar explanation may also hold for diseases of greater consequence such as breast cancer, for which an implied liability *R*^2^ of 16.1% better matches the sum of winner’s curse-unadjusted variance explained by all GWAS susceptibility loci (18%) in Michailidou et al. ^6^ than our concordant literature and UKBB validation estimates of liability *R*^2^ ∼ 9.5% (**Supplementary Note Table 2**). The same is true of the implied liability *R*^2^ versus SNP heritability for asthma (10.7% vs. 10.6% for adult-onset asthma ^7^), insomnia (10.3% vs. 9–11%, see the “SNP heritability” section of Hammerschlag et al. ^8^) and hypertension (11.7% vs. 10.7% ^9^), perhaps among others.

Given these cumulative concerns and lack of published external validation, we concluded that inclusion of Nucleus’ scores in our primary benchmarking comparison would not yield a reliable or meaningful addition to our analysis.

## Supplementary Tables

**Supplementary Table 1:**
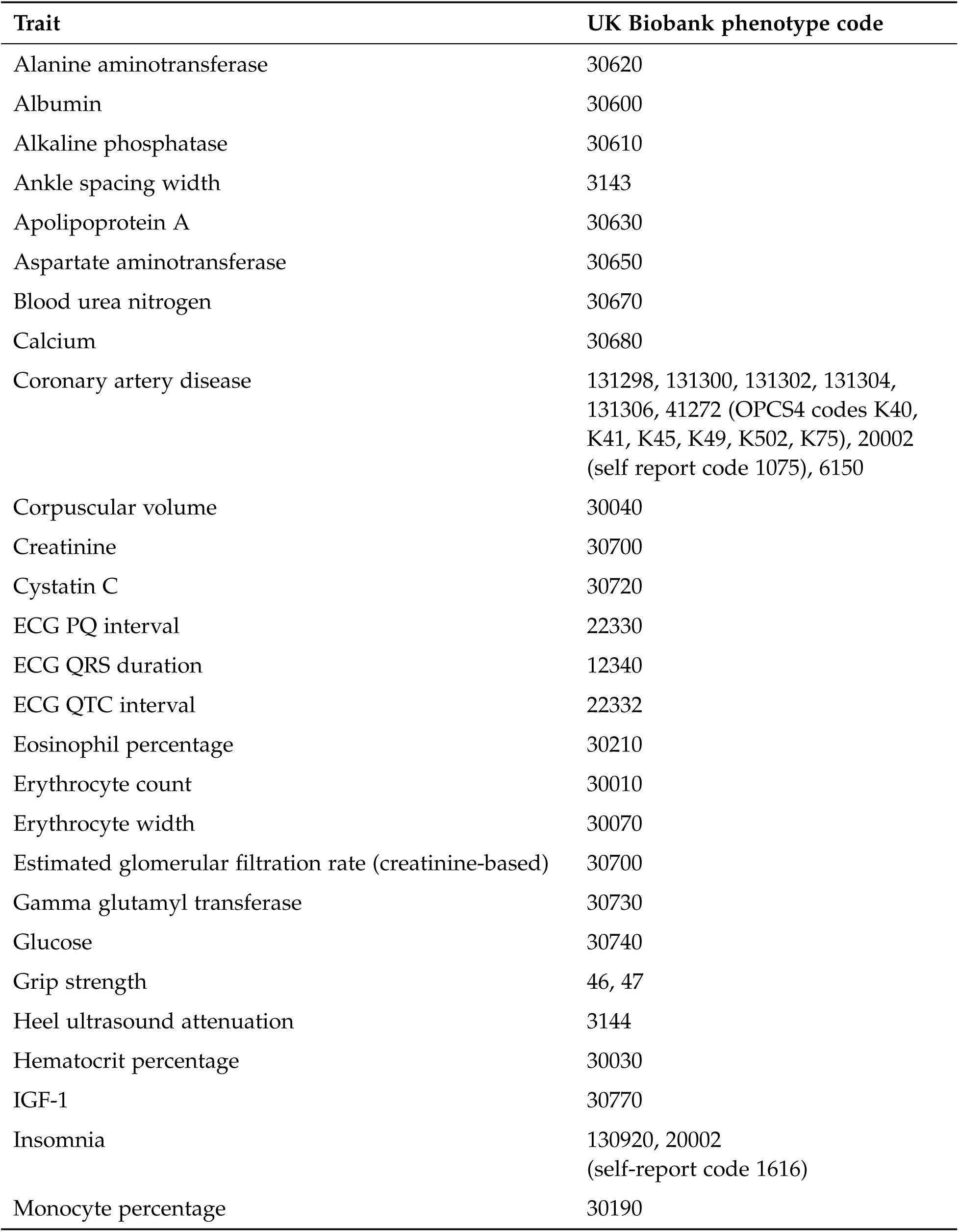
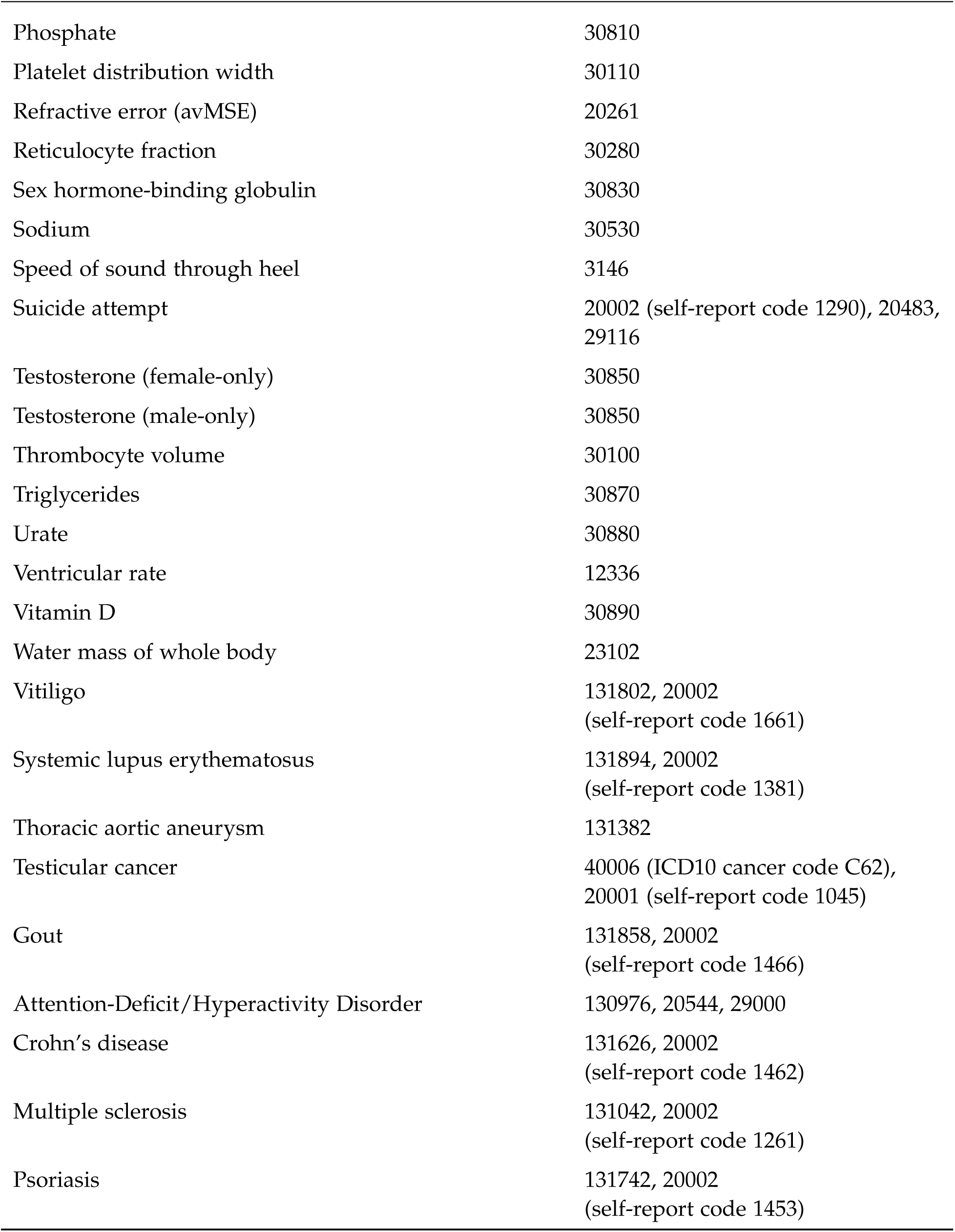
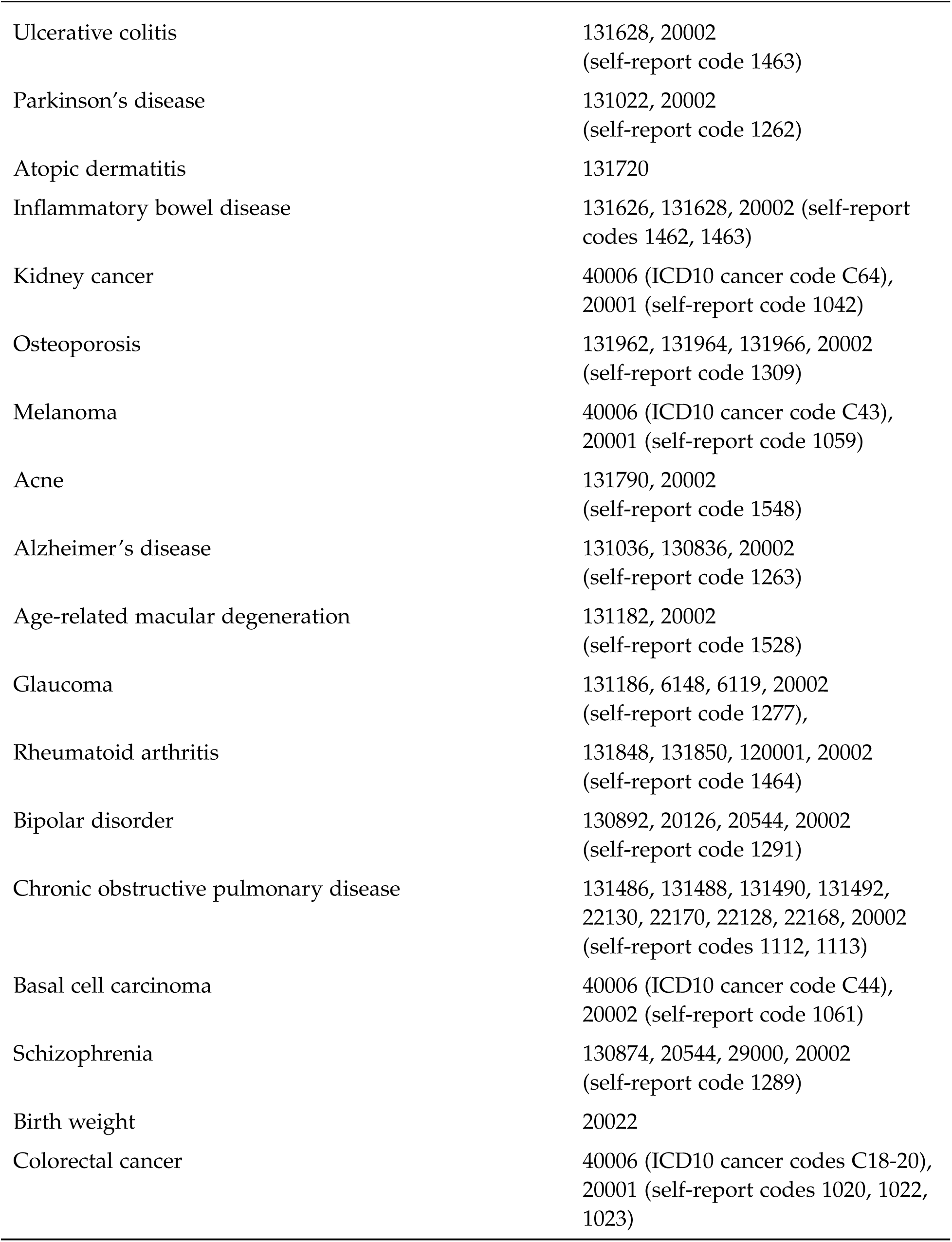
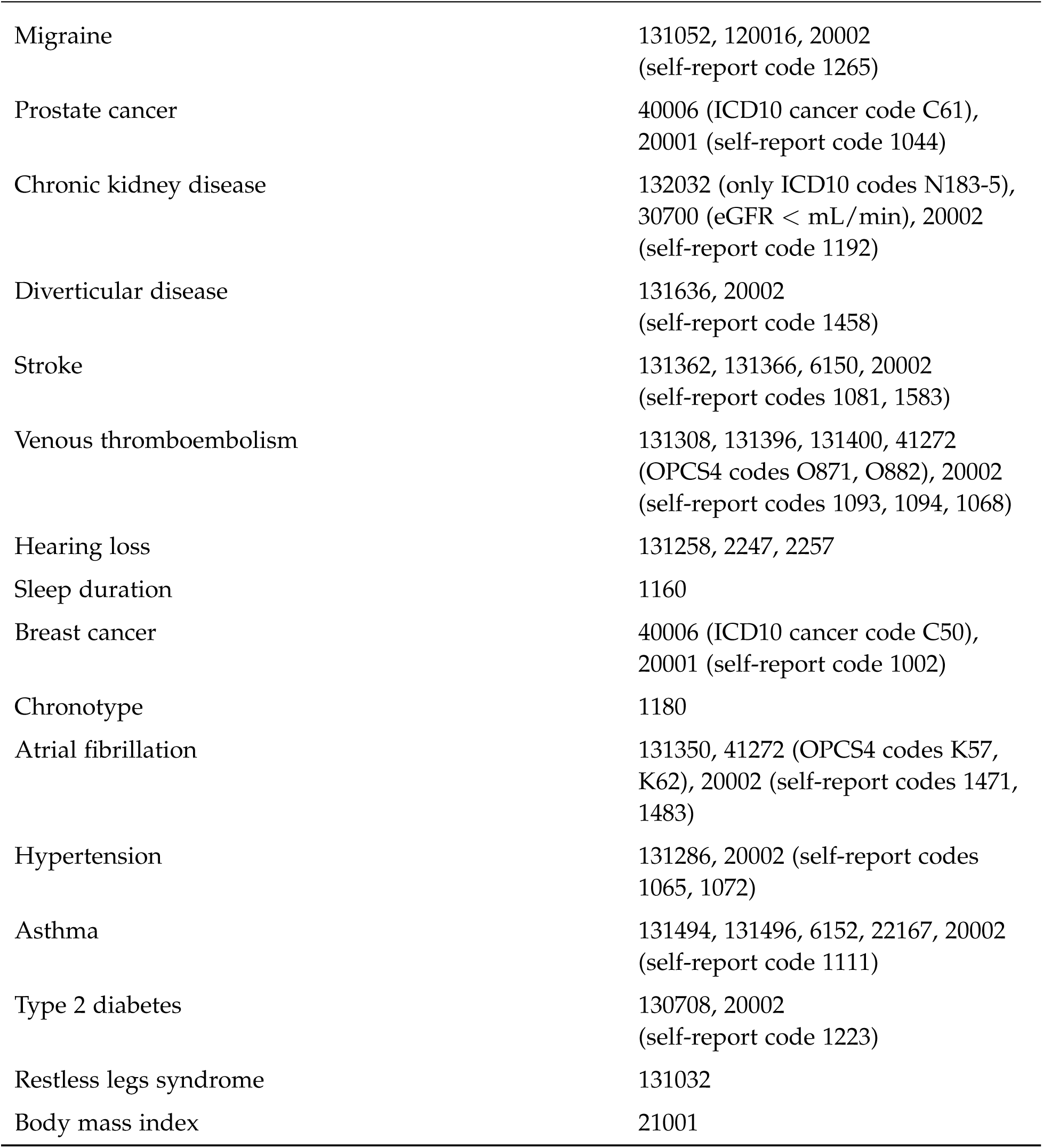
Phenotype definitions used for disease and the traits used in UK Biobank for validation and relative PGS performance attenuation in diverse ancestries.

| Trait | UK Biobank phenotype code |
| --- | --- |
| Alanine aminotransferase | 30620 |
| Albumin | 30600 |
| Alkaline phosphatase | 30610 |
| Ankle spacing width | 3143 |
| Apolipoprotein A | 30630 |
| Aspartate aminotransferase | 30650 |
| Blood urea nitrogen | 30670 |
| Calcium | 30680 |
| Coronary artery disease | 131298, 131300, 131302, 131304, 131306, 41272 (OPCS4 codes K40, K41, K45, K49, K502, K75), 20002 (self report code 1075), 6150 |
| Corpuscular volume | 30040 |
| Creatinine | 30700 |
| Cystatin C | 30720 |
| ECG PQ interval | 22330 |
| ECG QRS duration | 12340 |
| ECG QTC interval | 22332 |
| Eosinophil percentage | 30210 |
| Erythrocyte count | 30010 |
| Erythrocyte width | 30070 |
| Estimated glomerular filtration rate (creatinine-based) | 30700 |
| Gamma glutamyl transferase | 30730 |
| Glucose | 30740 |
| Grip strength | 46, 47 |
| Heel ultrasound attenuation | 3144 |
| Hematocrit percentage | 30030 |
| IGF-1 | 30770 |
| Insomnia | 130920, 20002 (self-report code 1616) |
| Monocyte percentage | 30190 |

| **Trait** | **UK Biobank phenotype code** |
| --- | --- |
| Phosphate | 30810 |
| Platelet distribution width | 30110 |
| Refractive error (avMSE) | 20261 |
| Reticulocyte fraction | 30280 |
| Sex hormone-binding globulin | 30830 |
| Sodium | 30530 |
| Speed of sound through heel | 3146 |
| Suicide attempt | 20002 (self-report code 1290), 20483, 29116 |
| Testosterone (female-only) | 30850 |
| Testosterone (male-only) | 30850 |
| Thrombocyte volume | 30100 |
| Triglycerides | 30870 |
| Urate | 30880 |
| Ventricular rate | 12336 |
| Vitamin D | 30890 |
| Water mass of whole body | 23102 |
| Vitiligo | 131802, 20002<br>(self-report code 1661) |
| Systemic lupus erythematosus | 131894, 20002<br>(self-report code 1381) |
| Thoracic aortic aneurysm | 131382 |
| Testicular cancer | 40006 (ICD10 cancer code C62),<br>20001 (self-report code 1045) |
| Gout | 131858, 20002<br>(self-report code 1466) |
| Attention-Deficit/Hyperactivity Disorder | 130976, 20544, 29000 |
| Crohn's disease | 131626, 20002<br>(self-report code 1462) |
| Multiple sclerosis | 131042, 20002<br>(self-report code 1261) |
| Psoriasis | 131742, 20002<br>(self-report code 1453) |
| Ulcerative colitis | 131628, 20002<br>(self-report code 1463) |
| Parkinson's disease | 131022, 20002<br>(self-report code 1262) |
| Atopic dermatitis | 131720 |
| Inflammatory bowel disease | 131626, 131628, 20002 (self-report codes 1462, 1463) |
| Kidney cancer | 40006 (ICD10 cancer code C64),<br>20001 (self-report code 1042) |
| Osteoporosis | 131962, 131964, 131966, 20002<br>(self-report code 1309) |
| Melanoma | 40006 (ICD10 cancer code C43),<br>20001 (self-report code 1059) |
| Acne | 131790, 20002<br>(self-report code 1548) |
| Alzheimer's disease | 131036, 130836, 20002<br>(self-report code 1263) |
| Age-related macular degeneration | 131182, 20002<br>(self-report code 1528) |
| Glaucoma | 131186, 6148, 6119, 20002<br>(self-report code 1277), |
| Rheumatoid arthritis | 131848, 131850, 120001, 20002<br>(self-report code 1464) |
| Bipolar disorder | 130892, 20126, 20544, 20002<br>(self-report code 1291) |
| Chronic obstructive pulmonary disease | 131486, 131488, 131490, 131492,<br>22130, 22170, 22128, 22168, 20002<br>(self-report codes 1112, 1113) |
| Basal cell carcinoma | 40006 (ICD10 cancer code C44),<br>20002 (self-report code 1061) |
| Schizophrenia | 130874, 20544, 29000, 20002<br>(self-report code 1289) |
| Birth weight | 20022 |
| Colorectal cancer | 40006 (ICD10 cancer codes C18-20),<br>20001 (self-report codes 1020, 1022,<br>1023) |
| Migraine | 131052, 120016, 20002<br>(self-report code 1265) |
| Prostate cancer | 40006 (ICD10 cancer code C61),<br>20001 (self-report code 1044) |
| Chronic kidney disease | 132032 (only ICD10 codes N183-5),<br>30700 (eGFR < mL/min), 20002<br>(self-report code 1192) |
| Diverticular disease | 131636, 20002<br>(self-report code 1458) |
| Stroke | 131362, 131366, 6150, 20002<br>(self-report codes 1081, 1583) |
| Venous thromboembolism | 131308, 131396, 131400, 41272<br>(OPCS4 codes O871, O882), 20002<br>(self-report codes 1093, 1094, 1068) |
| Hearing loss | 131258, 2247, 2257 |
| Sleep duration | 1160 |
| Breast cancer | 40006 (ICD10 cancer code C50),<br>20001 (self-report code 1002) |
| Chronotype | 1180 |
| Atrial fibrillation | 131350, 41272 (OPCS4 codes K57,<br>K62), 20002 (self-report codes 1471,<br>1483) |
| Hypertension | 131286, 20002 (self-report codes<br>1065, 1072) |
| Asthma | 131494, 131496, 6152, 22167, 20002<br>(self-report code 1111) |
| Type 2 diabetes | 130708, 20002<br>(self-report code 1223) |
| Restless legs syndrome | 131032 |
| Body mass index | 21001 |

**Supplementary Table 2:**
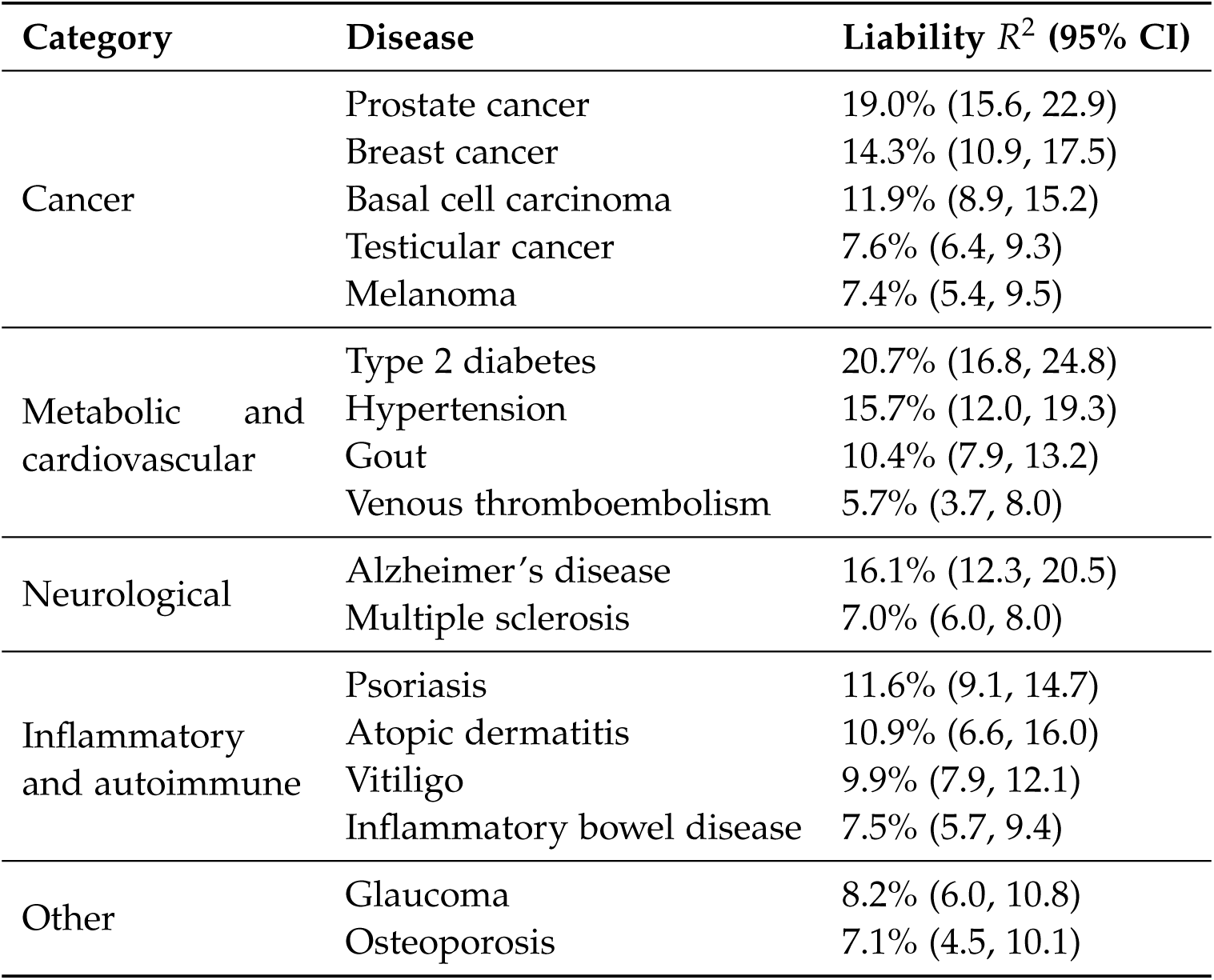
Liability *R*^2^ estimated between families in the UK Biobank for 17 newly developed disease PGS.

| Category | Disease | Liability $R^2$ (95% CI) |
| --- | --- | --- |
| Cancer | Prostate cancer | 19.0% (15.6, 22.9) |
|  | Breast cancer | 14.3% (10.9, 17.5) |
|  | Basal cell carcinoma | 11.9% (8.9, 15.2) |
|  | Testicular cancer | 7.6% (6.4, 9.3) |
|  | Melanoma | 7.4% (5.4, 9.5) |
| Metabolic and cardiovascular | Type 2 diabetes | 20.7% (16.8, 24.8) |
|  | Hypertension | 15.7% (12.0, 19.3) |
|  | Gout | 10.4% (7.9, 13.2) |
|  | Venous thromboembolism | 5.7% (3.7, 8.0) |
| Neurological | Alzheimer's disease | 16.1% (12.3, 20.5) |
|  | Multiple sclerosis | 7.0% (6.0, 8.0) |
| Inflammatory and autoimmune | Psoriasis | 11.6% (9.1, 14.7) |
|  | Atopic dermatitis | 10.9% (6.6, 16.0) |
|  | Vitiligo | 9.9% (7.9, 12.1) |
|  | Inflammatory bowel disease | 7.5% (5.7, 9.4) |
| Other | Glaucoma | 8.2% (6.0, 10.8) |
|  | Osteoporosis | 7.1% (4.5, 10.1) |

**Supplementary Table 3:** Lifetime risk estimates and their corresponding sources for European ancestry individuals for males (M) and females (F).

| Disease | Lifetime risk (EUR) |  | Source |  |
| --- | --- | --- | --- | --- |
|  | M | F |  |  |
| Vitiligo | 0.45% | 0.46% | Howitz et al., 1977 | <sup>10</sup> |
| Testicular cancer | 0.49% |  | SEER, 2024 | <sup>11</sup> |
| Gout | 11.00% | 3.10% | Dehlin et al., 2020 | <sup>12</sup> |
| Multiple sclerosis | 0.19% | 0.54% | Hittle et al. 2023 | <sup>13</sup> |
| Psoriasis | 2.90% | 2.90% | Pezzolo et al., 2019 | <sup>14</sup> |
| Atopic dermatitis | 12.00% | 12.00% | Hadi et al., 2021 | <sup>15</sup> |
| Inflammatory bowel disease | 1.69% | 1.85% | Forss et al. 2022 | <sup>16</sup> |
| Osteoporosis | 22.00% | 46.00% | Kanis et al., 2000 | <sup>17</sup> |
| Melanoma | 3.47% | 2.48% | SEER, 2024 | <sup>11</sup> |
| Alzheimer's disease | 8.80% | 13.40% | Lobo et al., 2011 | <sup>18</sup> |
| Glaucoma | 5.53% | 8.51% | NEI, 2024 | <sup>19</sup> |
| Venous thromboembolism | 7.70% | 8.40% | Bell et al., 2016 | <sup>20</sup> |
| Prostate cancer | 12.57% |  | SEER, 2024 | <sup>11</sup> |
| Basal cell carcinoma | 21.00% | 18.00% | Flohil et al., 2013 | <sup>21</sup> |
| Breast cancer |  | 13.87% | SEER, 2024 | <sup>11</sup> |
| Hypertension | 45.40% | 48.30% | FinnGen, 2024 | <sup>22</sup> |
| Type 2 diabetes | 21.40% | 18.40% | Narayan et al., 2007 | <sup>23</sup> |

**Supplementary Table 4:** Lifetime risk estimates for type 2 diabetes by individual ancestries as reported in Narayan et al. (2007) ^23^ for males (M) and females (F).

| Ancestry | Type 2 diabetes lifetime risk |  |
| --- | --- | --- |
|  | M | F |
| SAS | 26.40% | 24.19% |
| EAS | 27.59% | 23.72% |
| AFR | 26.35% | 28.61% |

**Supplementary Table 5:**
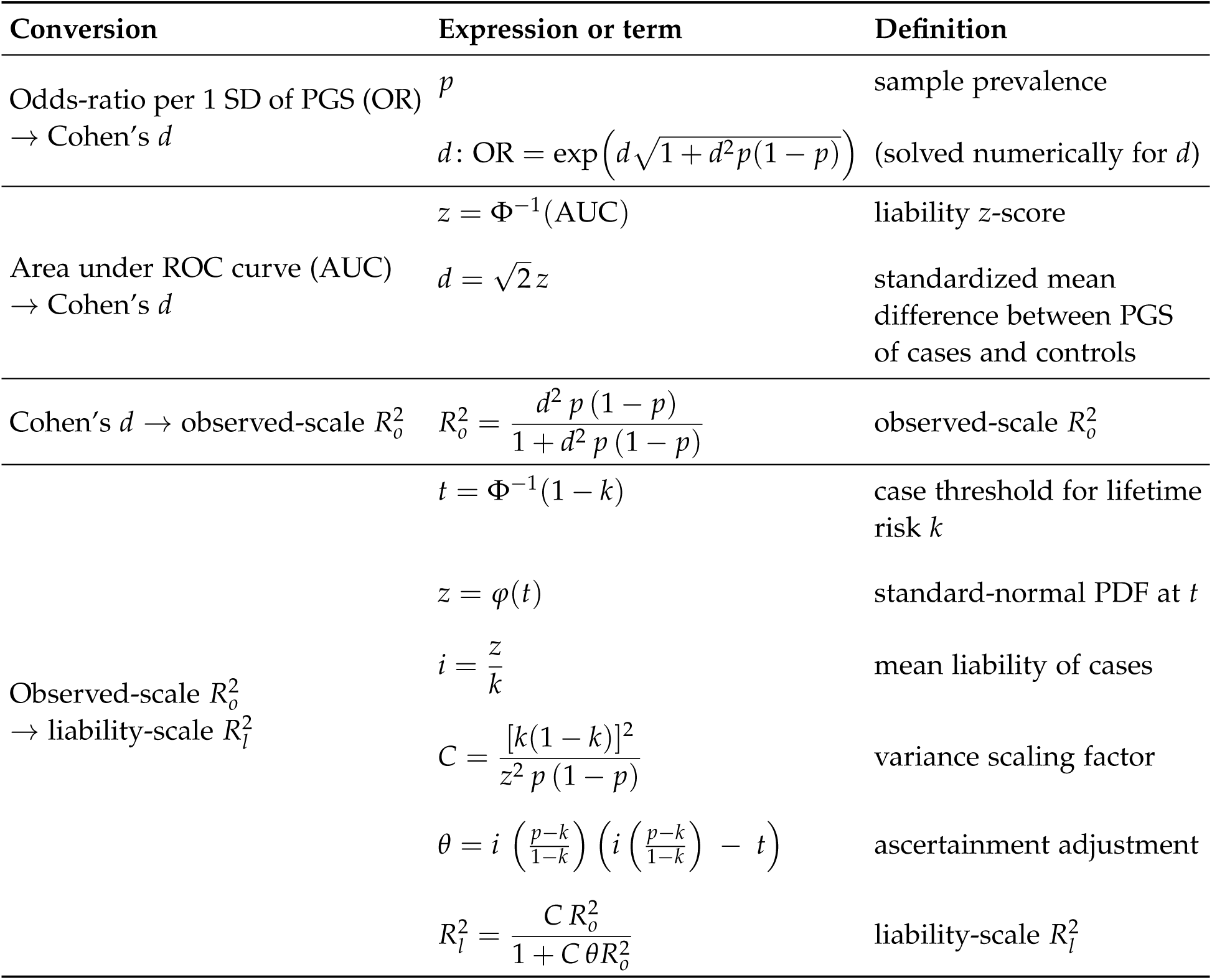
Equations used to convert comparison validations reported in odds ratios or AUC to liability *R*^2^.

**Supplementary Table 6:**
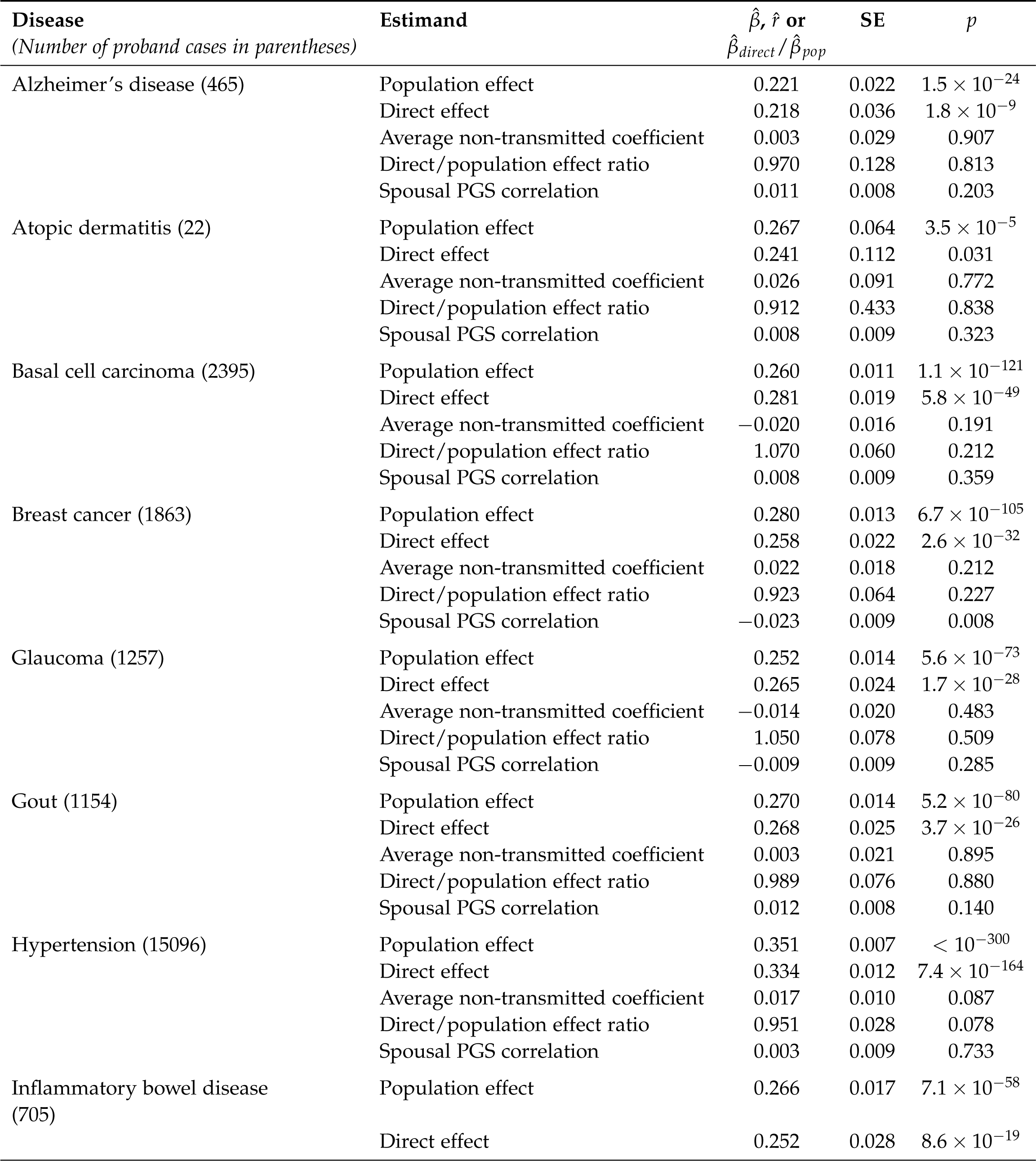
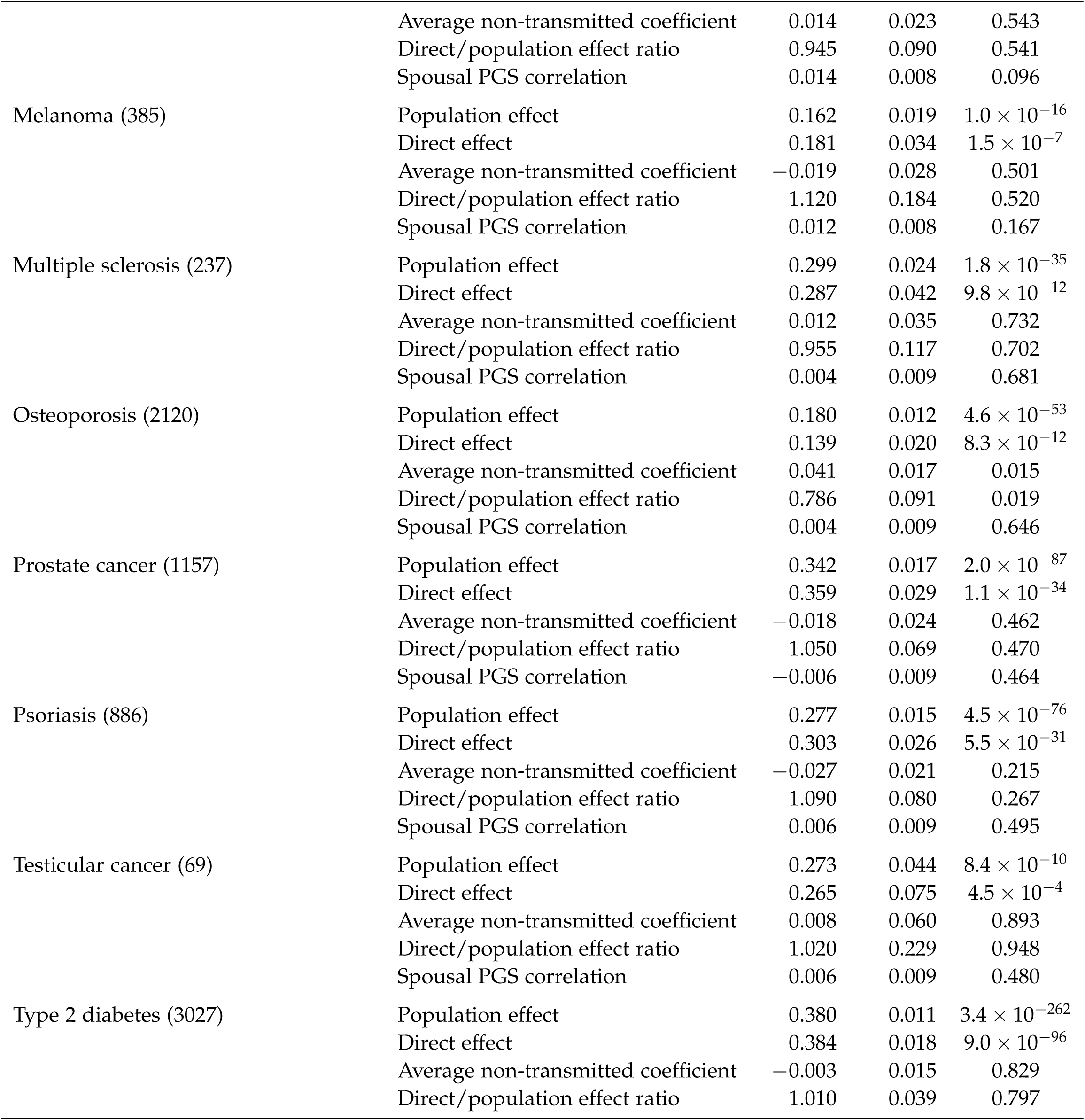
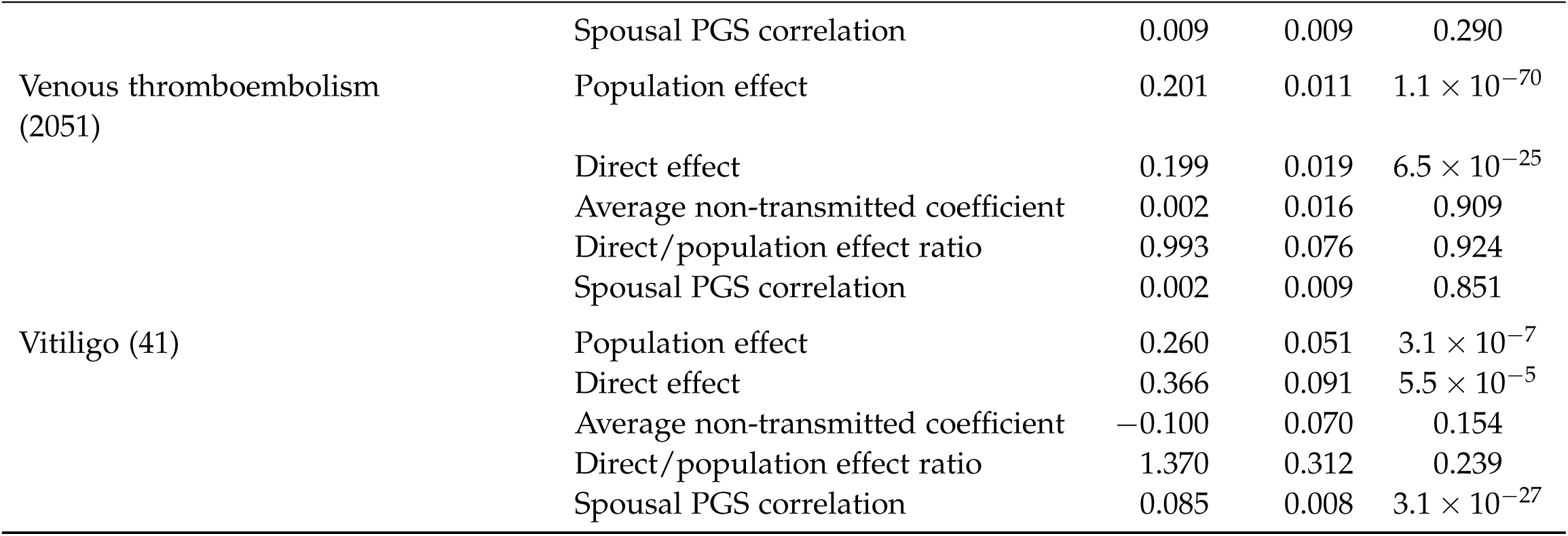
Within-family model estimates. “Direct effect” and “Average non-transmitted coefficient” refer to the estimated coefficients on a proband PGS term and combined maternal + paternal PGS term in the within-family specification, respectively. “Direct/population effect ratio” refers to the ratio of direct to population effects derived from a within-family model fitting maternal and paternal PGS terms separately.

**Supplementary Table 7:** Sample prevalences used to convert PGS AUC reported by commercial PGT-P providers to liability *R*^2^ shown in Figure 3. Values for Genomic Prediction were approximated using the available information in the supplement to Widen et al. ^24^ and on Genomic Prediction’s risk reduction calculator webpage ^25^. Values for Orchid were found in their online whitepapers ^26^.

| Disease | Orchid sample prevalence | Genomic Prediction sample prevalence |
| --- | --- | --- |
| Alzheimer's | omitted | ~1.2% |
| Basal cell carcinoma | not validated | ~1.4% |
| Breast cancer | 8.0% | ~4.7% |
| Type 2 diabetes | 7.5% | ~6.4% |
| Gout | not validated | ~2% |
| Hypertension | not validated | ~32% |
| Inflammatory bowel disease | 1.5% | ~1.5% |
| Melanoma | not validated | ~0.9% |
| Prostate cancer | 7.4% | ~2.1% |
| Testicular cancer | not validated | ~0.4% |

**Supplementary Table 8:**
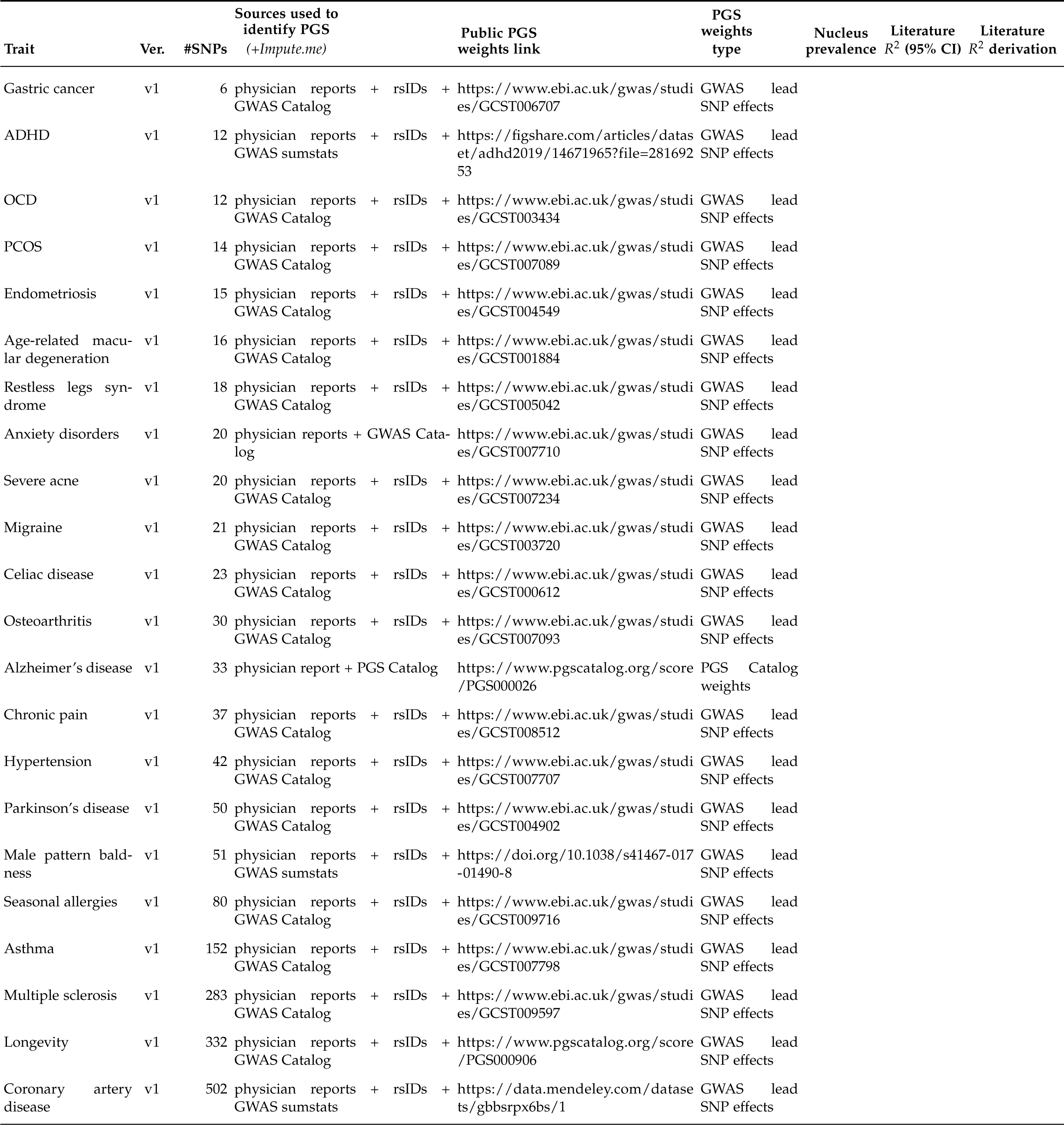
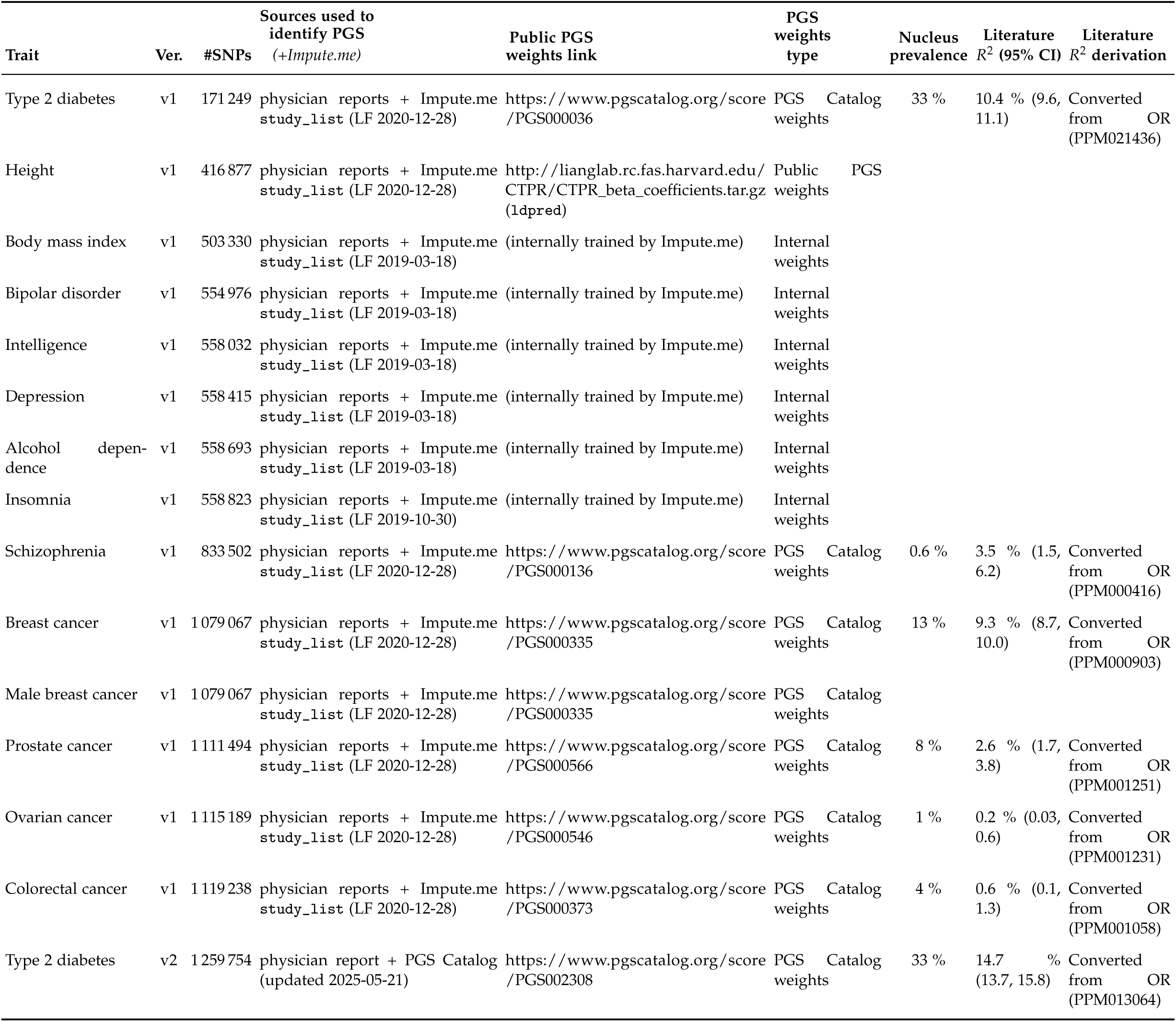
Nucleus PGS models reconstructed by matching variant counts shown on physician reports with external sources of SNP rsIDs and weights. “Impute.me study_list sheet” refers to 2021-02-11_study_list.xlsx on the Impute.me GitHub repository. ^27^

**Supplementary Table 9:** Summary statistics of liability *R*^2^ per trait, including the median from European-ancestry reports.

| Disease | N | Minimum | q25 | Median | Median<br>(EUR<br>reports<br>only) | Mean | q75 | Maximum | SD |
| --- | --- | --- | --- | --- | --- | --- | --- | --- | --- |
| Breast cancer | 1 | 0.1613 | 0.1613 | 0.1613 | 0.1613 | 0.1613 | 0.1613 | 0.1613 | NA |
| Migraine | 21 | 0.0004 | 0.0741 | 0.1525 | 0.1764 | 0.1252 | 0.1949 | 0.2181 | 0.0768 |
| Coronary artery disease | 13 | 0.0226 | 0.0835 | 0.1187 | 0.1522 | 0.1184 | 0.1570 | 0.1734 | 0.0462 |
| Type 2 diabetes | 21 | 0.0374 | 0.0850 | 0.1220 | 0.1306 | 0.1143 | 0.1367 | 0.2718 | 0.0498 |
| Chronic pain | 16 | 0.0489 | 0.0854 | 0.1160 | 0.1319 | 0.1132 | 0.1420 | 0.1592 | 0.0359 |
| Parkinson's disease | 5 | 0.0170 | 0.0787 | 0.1311 | 0.1392 | 0.1068 | 0.1392 | 0.1681 | 0.0597 |
| Restless legs syndrome | 5 | 0.0616 | 0.0806 | 0.0942 | 0.0942 | 0.0942 | 0.1172 | 0.1174 | 0.0241 |
| Asthma | 12 | 0.0468 | 0.0802 | 0.0993 | 0.1073 | 0.0937 | 0.1144 | 0.1341 | 0.0294 |
| Insomnia | 16 | 0.0461 | 0.0776 | 0.0989 | 0.1034 | 0.0923 | 0.1131 | 0.1280 | 0.0272 |
| Endometriosis | 1 | 0.0890 | 0.0890 | 0.0890 | 0.0890 | 0.0890 | 0.0890 | 0.0890 | NA |
| Hypertension | 17 | 0.0038 | 0.0617 | 0.1065 | 0.1173 | 0.0886 | 0.1268 | 0.1407 | 0.0469 |
| Alcohol dependence | 8 | 0.0004 | 0.0533 | 0.0899 | 0.1063 | 0.0839 | 0.1153 | 0.1514 | 0.0478 |
| PCOS | 3 | 0.0803 | 0.0808 | 0.0812 | 0.0812 | 0.0830 | 0.0843 | 0.0874 | 0.0038 |
| Male breast cancer | 2 | 0.0693 | 0.0748 | 0.0803 | 0.0803 | 0.0803 | 0.0858 | 0.0913 | 0.0155 |
| Multiple sclerosis | 5 | 0.0128 | 0.0437 | 0.1003 | 0.1023 | 0.0777 | 0.1023 | 0.1294 | 0.0479 |
| OCD | 4 | 0.0486 | 0.0517 | 0.0739 | 0.0950 | 0.0776 | 0.0998 | 0.1142 | 0.0322 |
| Age-related macular<br>degeneration | 20 | 0.0019 | 0.0404 | 0.0840 | 0.0925 | 0.0767 | 0.1118 | 0.1479 | 0.0463 |
| Prostate cancer | 13 | 0.0220 | 0.0442 | 0.0697 | 0.0744 | 0.0746 | 0.0898 | 0.2158 | 0.0494 |
| Anxiety disorders | 16 | 0.0332 | 0.0412 | 0.0753 | 0.0827 | 0.0698 | 0.0852 | 0.1036 | 0.0246 |
| Rheumatoid arthritis | 20 | 0.0000 | 0.0495 | 0.0773 | 0.0782 | 0.0676 | 0.0895 | 0.1291 | 0.0358 |
| Severe acne | 3 | 0.0145 | 0.0237 | 0.0328 | 0.0812 | 0.0595 | 0.0820 | 0.1311 | 0.0627 |
| ADHD | 13 | 0.0181 | 0.0286 | 0.0405 | 0.0429 | 0.0408 | 0.0509 | 0.0716 | 0.0149 |
| Schizophrenia | 24 | 0.0000 | 0.0241 | 0.0312 | 0.0372 | 0.0317 | 0.0410 | 0.0499 | 0.0137 |
| Colorectal cancer | 5 | 0.0124 | 0.0253 | 0.0254 | 0.0270 | 0.0236 | 0.0270 | 0.0280 | 0.0064 |
| Osteoarthritis | 12 | 0.0090 | 0.0158 | 0.0236 | 0.0312 | 0.0236 | 0.0321 | 0.0344 | 0.0095 |
| Bipolar disorder | 5 | 0.0072 | 0.0110 | 0.0111 | 0.0123 | 0.0108 | 0.0123 | 0.0124 | 0.0021 |
| Depression | 4 | 0.0024 | 0.0040 | 0.0051 | 0.0057 | 0.0107 | 0.0118 | 0.0301 | 0.0130 |
| Ovarian cancer | 1 | 0.0010 | 0.0010 | 0.0010 | 0.0011 | 0.0010 | 0.0010 | 0.0010 | NA |
| Gastric cancer | 5 | 0.0000 | 0.0000 | 0.0008 | 0.0009 | 0.0010 | 0.0014 | 0.0026 | 0.0011 |

**Supplementary Table 10:**
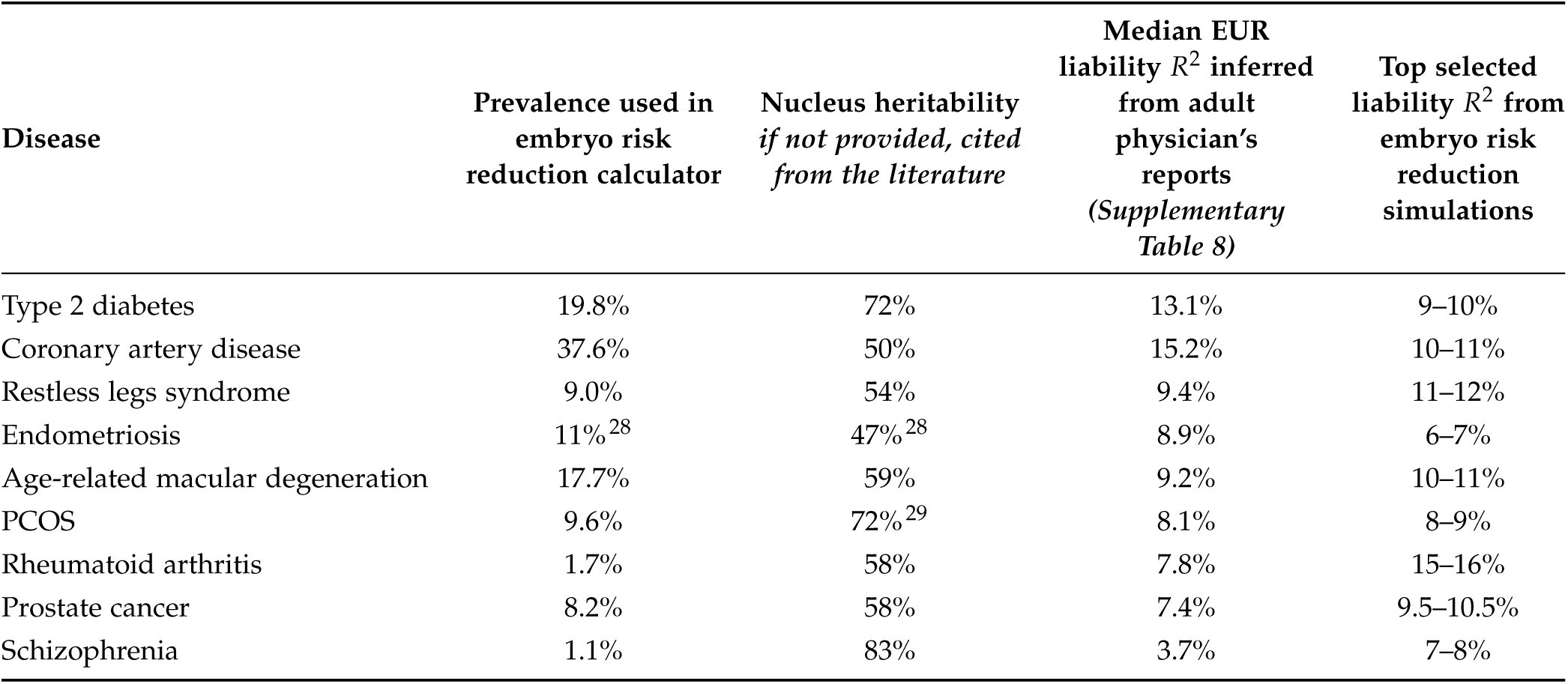
Liability *R*^2^ inferred from Nucleus’ adult physician’s reports and embryo risk-reduction calculator ^1^. Prevalences and heritabilities were scraped from Nucleus materials for input to risk-reduction simulations unless otherwise noted.

| Disease | Prevalence used in embryo risk reduction calculator | Nucleus heritability if not provided, cited from the literature | Median EUR liability $R^2$ inferred from adult physician's reports (Supplementary Table 8) | Top selected liability $R^2$ from embryo risk reduction simulations |
| --- | --- | --- | --- | --- |
| Type 2 diabetes | 19.8% | 72% | 13.1% | 9–10% |
| Coronary artery disease | 37.6% | 50% | 15.2% | 10–11% |
| Restless legs syndrome | 9.0% | 54% | 9.4% | 11–12% |
| Endometriosis | 11%<sup>28</sup> | 47%<sup>28</sup> | 8.9% | 6–7% |
| Age-related macular degeneration | 17.7% | 59% | 9.2% | 10–11% |
| PCOS | 9.6% | 72%<sup>29</sup> | 8.1% | 8–9% |
| Rheumatoid arthritis | 1.7% | 58% | 7.8% | 15–16% |
| Prostate cancer | 8.2% | 58% | 7.4% | 9.5–10.5% |
| Schizophrenia | 1.1% | 83% | 3.7% | 7–8% |

